# Estimating age-specific heterogeneity in SARS-CoV-2 transmission from prospective longitudinal studies: the importance of correcting for study design

**DOI:** 10.64898/2026.08.06.26358866

**Authors:** Sophie Chervet, Maylis Layan, Pierre-Yves Boëlle, Jérémie Guedj, Sylvie van der Werf, Solen Kernéis, Isabelle Sermet-Gaudelus, Simon Cauchemez, Lulla Opatowski

**Author notes:** These authors contributed equally (Isabelle Sermet-Gaudelus, Simon Cauchemez, Lulla Opatowski).

## Abstract

Longitudinal household studies, combined with mathematical modeling, are widely used to characterize the drivers of respiratory pathogen transmission, including the effects of age and symptoms. In practice, household recruitment protocols vary across studies, potentially introducing biases into observed data. However, these biases are typically overlooked in statistical inference, and their impact on parameter estimates remains unknown. Here, we use synthetic household outbreak data simulated under different recruitment protocols to evaluate how recruiting through infected children affects estimates of age-specific infectiousness and susceptibility. We show that, under child-based recruitment, the standard likelihood, which accounts only for transmission dynamics, leads to underestimating child infectiousness and overestimating child susceptibility by more than 30%. We then propose a novel estimation framework that explicitly incorporates the household recruitment process into the likelihood and show that it substantially reduces these biases. Applying this new approach to a French household study conducted during the COVID-19 pandemic, we estimated that children under 6 had 49% lower infectiousness than teenagers and adults during the Alpha wave, whereas no difference was observed during the Omicron wave. This study demonstrates that ignoring recruitment protocols can bias key epidemiological parameter estimates and highlights the importance of accounting for study design.

## Introduction

Household studies are important tools for investigating the transmission of respiratory viruses. This is because the high density of household contacts favors transmission. Consequently, household studies have been used to explore key questions about transmission drivers^1,2^. For example, during the SARS-CoV-2 pandemic, several household studies examined the role of children in SARS-CoV-2 transmission^2–7^, which is key to evaluate the impact of non-pharmaceutical interventions such as school closures. Children were rapidly considered less at risk of both infection and transmission for SARS-CoV-2, in contrast to their large role in influenza transmission^8,9^. However, many studies did not distinguish between age groups within the child population (e.g. pre-school vs school-aged children) although their transmission characteristics may differ^2,10^.

A common design for household studies consists in following up a household for a few weeks after the identification of a case (the inclusion case) in the household. Modeling is necessary to address key challenges for the reconstruction of household transmission dynamics after recruitment, e.g. the fact that household members may be infected by different sources in or out of the household, or that dates of infection are unknown^6,8,9,11^. However, little attention has been given to the way the strategy for household recruitment may affect parameter estimates. A simple strategy consists in identifying inclusion cases in patients attending general practice. However, this may provide limited power if the aim is to better understand the contribution of children to transmission. Over-representing children by recruiting inclusion cases in pediatric populations may address this power issue; but it can potentially bias parameter estimates overall. For example, if the first case of a household is an adult, this household can only be recruited if the adult case infects at least one child, pushing estimates of adult infectiousness upward.

Here, we show how recruiting households through infected children can bias estimates of the contribution of age groups to transmission. We then develop an inference framework that explicitly accounts for the recruitment protocol to overcome this bias. We use this framework to investigate the transmission dynamics of two SARS-CoV-2 variants of concern (Alpha and Omicron) from a prospective longitudinal study of French households recruited through positive children and explore the role of age in transmission.

## Methods

### PedCovid: a household study with recruitment through children

PedCovid (PI: I. Sermet-Gaudelus) is a prospective longitudinal study that took place in France with two independent recruitment periods^12,13^. The first period ran from March 11, 2021, to June 10, 2021, while the Alpha variant was predominantly circulating in France^14^ (“Alpha period”). The second period ran from December 14, 2021, to February 17, 2022, while the Omicron variant was predominant in France^14^ (“Omicron period”). The design of the PedCovid study is as follows: cases aged 18 and under (hereafter referred to as “inclusion cases”) were recruited at school after identification by a positive reverse transcription polymerase chain reaction (RT-PCR) test, either following symptoms or as contact cases during national screening campaigns. All household members who agreed to participate were then enrolled.

On the day of inclusion (D0), a saliva sample for the detection of SARS-CoV-2 by RT-PCR was collected from the inclusion case and all household members, as well as a serologic sample and a nasopharyngeal sample if accepted. All participants were then followed up for 45 days, with four visits: on day 3 (D3), day 7 (D7), day 15 (D15) and day 45 (D45) after inclusion. At each visit, saliva samples were collected. Demographic and clinical data were collected, including age, gender, vaccination status, previous SARS-CoV-2 infection, clinical status (symptomatic or asymptomatic), date and type of symptoms if applicable, and all dates and results of tests during the study period, whether during our outside visits. Data also included basic information on the control measures applied within households, along with the end date of implementation for each individual. In particular, households were asked whether they quarantined infected individuals in an isolated room (“isolation”), and whether they disinfected surfaces on a daily basis (“disinfection”).

This study protocol was approved by the ethics committee of Sud-Méditerranée V (n°20.04.14.62339) and was registered (clinicaltrials.gov identifier NCT04355533).

To analyze the effect of age, we divided the population into three age categories based on school ages in France: “infants” (younger than 6 years old), “children” (from 6 to 11 years old) and “teenagers and adults” (older than 11 years old).

### Transmission model

An individual-based model of household transmission was developed and integrated within an inference framework. The model was extended from Layan et al^6^.

The model allows for individuals to avoid infection, be infected in the community, or be infected by household members. It takes into account individual characteristics such as age, symptoms and past exposure from vaccination or past infection; household characteristics such as household size and control measures implemented; and virus characteristics through the distributions of the incubation period (time between infection and symptoms) and the generation time (time between the infections of the infector and their secondary case).

At time *t*, the instantaneous risk of infection of individual *i* in household *H*_*i*_, *λ*_*i*_ (*t*), is the sum of the risk of infection from the community *c* and the infection hazards from all other household members *j* that were infected before:

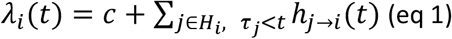

where *τ*_*j*_ is the day of infection of individual *j*. The community risk *c* is assumed constant over each study period. The instantaneous risk that infector *j* infects susceptible *I* at time *t, h*_*j*→*i*_ (*t*), is given by:

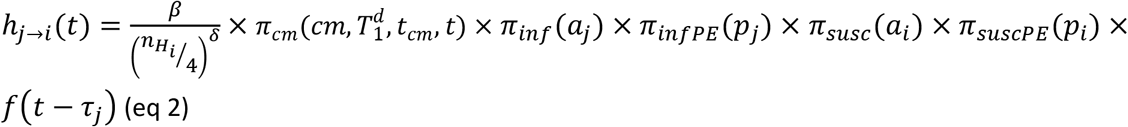

where *β* is the baseline person-to-person transmission intensity with a dependence on household size 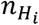 characterized by parameter *δ* (Figure 1). It is then modulated by:

**Figure 1.**
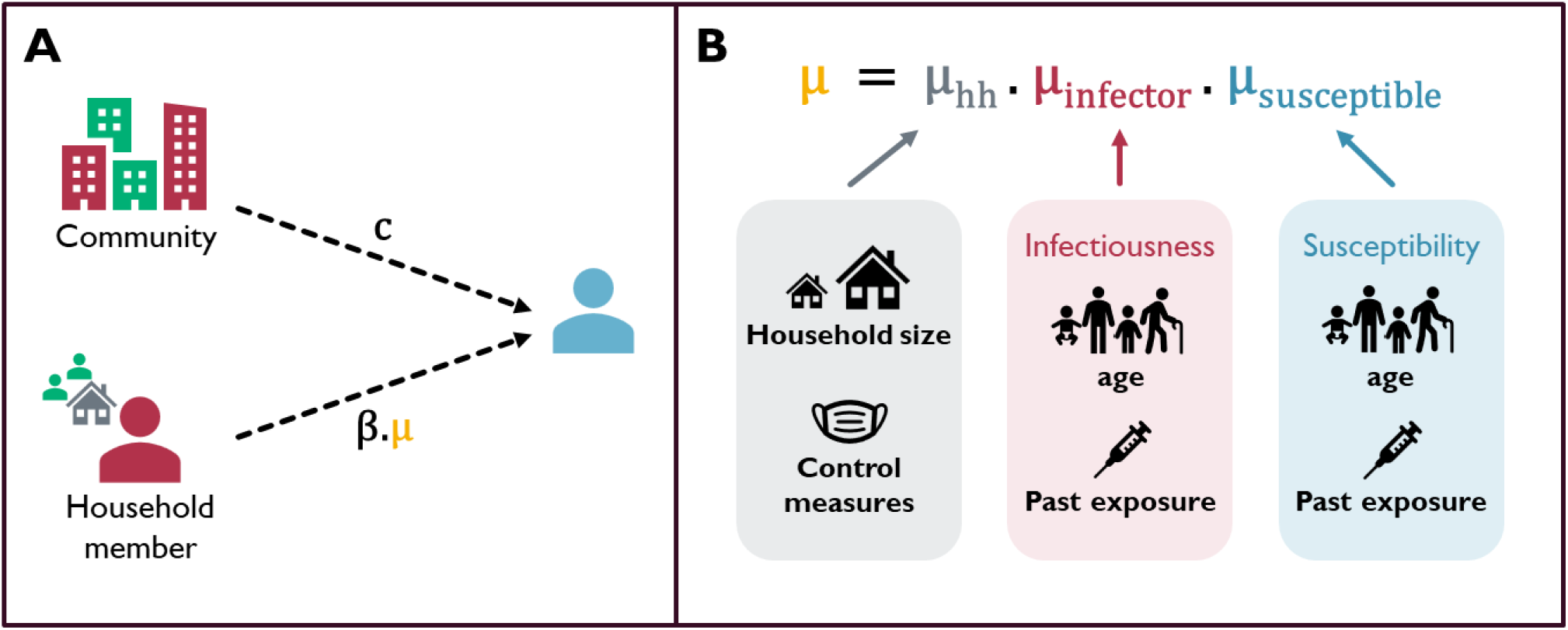
Schematic of our transmission model. A – Possible sources of infection for an individual and associated risk. c: risk of infection in the community, β: baseline inter-individual transmission intensity, μ: relative pairwise transmission factor. B – Decomposition of the relative pairwise transmission factor μ, with terms describing household characteristics μ_hh_,, individual characteristics of the infector μ_infector_, and individual characteristics of the susceptible μ_susceptible_ .

i. the relative infectiousness *π*_*inf*_ and *π*_*infPE*_ of the infector *j* according to their age *a*_*j*_ and past exposure *p*_*j*_ . *π*_*inf*_ equals 1 for teenagers/adults (reference), *µ*_*infc*_ for children, and *µ*_*infI*_ for infants. *π*_*infPE*_ equals 1 for not previously exposed (reference) and *µ*_*infPE*_ for previously exposed.
ii. the relative susceptibility *π*_*susc*_ and *π*_*suscPE*_ *o*f the susceptible *i*according to their age *a*_*i*_ and past exposure *p*_*i*_ . *π*_*susc*_ equals 1 for teenagers/adults (reference), *µ*_*suscc*_ for children, and *µ*_*suscI*_ for infants. *π*_*suscPE*_ equals 1 for not previously exposed (reference) and *µ*_*suscPE*_ for previously exposed.
iii. the effect *π*_*cm*_ of the control measures *cm* applied in the household at time *t*. Measures are applied between the first symptoms or positive test in the household 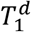 and the end of measures declared *t*_*cm*_ . *π*_*cm*_ equals 1 for no control measures (reference), *µ*_*cmDisinf*_ for disinfection of surfaces only, *µ*_*cmIsol*_ for household isolation only, and *µ*_*cmDisinf*_ × *µ*_*cmIsol*_ for both measures.
iv. the infectivity profile, i.e. the relative probability of transmission at time *t* given that the infector was infected at time *τ*_*j*_ . The profile *f* was informed by generation time distributions from the literature and is assumed to differ by variant (**Erreur ! Source du renvoi introuvable**.). The impact of the choice of distribution was evaluated in a sensitivity analysis (Supplementary Table 7).

### Inference framework

To estimate our parameters of interest, we used Bayesian Markov Chain Monte Carlo methods with data augmentation^8^. The observed data consist of symptom onset dates, and test dates and results for each individual. Data were augmented for each infected individual with the continuous-time dates of symptom onset/first positive test 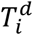and infection 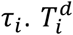 values were set within a day of the real observed date 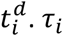 were constructed using the distribution *g* of the incubation period or duration of test positivity for asymptomatic cases.

All model parameters and distributions for both variants are summarized in Supplementary Table 1.

We use the Metropolis-Hastings algorithm to compute the posterior distribution of parameters *θ* and augmented data ***Z***. In case of random recruitment, this probability distribution is classically written:

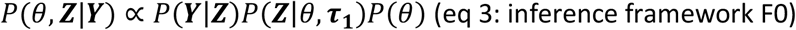

where ***Y*** are the observed data and 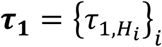 are the augmented infection dates of the index cases, i.e. the first infected in each household.

In the PedCovid study, households were recruited after detection of a positive child (≤18 years old). Here, we propose a new formulation that accounts for this nonrandom recruitment method by expressing the posterior probability as *P*(*θ*, ***Z***|***Y***, *R*), where *R* represents the recruitment method. The posterior probability develops as (see Supplementary section A.2.):

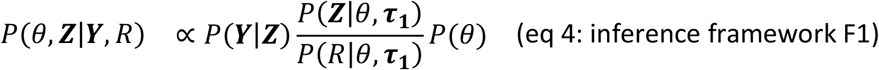

The likelihood of the transmission process *P*(***Z***|*θ, τ*_**1**_) and observation process *P*(***Y***|***Z***) classically used are described in Supplementary section A.3.

For the inference framework accounting for recruitment protocol (F1), we need to compute the likelihood of the recruitment constraint *P*(*R*|*θ, τ*_**1**_). Typically, a household would be recruited upon detection of an infected individual, the “inclusion individual”. However, in PedCovid, recruitment further requires that the inclusion individual is a child. We therefore computed *P*(*R*|*θ, τ*_**1**_) in each household as *P*(*inclusion individual is a child*| *inclusion individual is infected*), see Supplementary section A.3.

We compared the performance of the two inference frameworks (F0: classic likelihood, F1: corrected likelihood taking recruitment into account) in a simulation study.

#### Prior distributions and MCMC implementation

Prior distributions *P*(*θ*) are given in Supplementary Table 2 with a sensitivity analysis on the impact of the choice of priors presented in Supplementary Section C.8.b.

We ran chains for 300,000 iterations, recording one out of 500 iterations and discarding a burn-in period of 30,000 iterations. Convergence was assessed visually.

### Analysis of synthetic datasets with different recruitment protocols

#### Simulation of synthetic datasets

To evaluate the impact of the recruitment, we first analyzed synthetic datasets simulated using the individual-based model described above and a fixed set of parameters. In simulated datasets, households were enrolled following two recruitment protocols: a random recruitment where the inclusion case can be any household member with a positive test, and the recruitment protocol of PedCovid, i.e. where the inclusion case is a child (≤18 years old) with a positive test. Details on the simulation of outbreaks and of recruitments are provided in the Supplement (Supplementary Figures 1 and 2).

#### Comparison of inference frameworks

The two likelihood-based estimation frameworks (F0: classic likelihood, F1: corrected likelihood taking recruitment into account) were compared in their ability to recover the parameters from the simulated datasets with random and PedCovid-like recruitment protocols.

The null hypothesis, where we supposed homogeneous infectiousness and susceptibility across ages (all parameters equal to 1), was first evaluated. Baseline values for *c, β* and *δ* were defined as the median posterior estimates from the null model, i.e. the model where only *c, β* and *δ* are estimated (see Supplementary Table 6). Then, each parameter was varied (see Supplementary Table 4) while maintaining the others to baseline. Comparison between estimates and true value was done using two metrics: coverage, i.e. proportion of simulations for which the credible interval contains the true value, in percentage; and relative bias, i.e. (*posterior median* − *true value*) / *true value*, and its mean across simulations.

Finally, we performed a multivariate validation of framework F1 by using 200 independent datasets simulated with parameter values drawn from the posterior distribution obtained from the PedCovid data (Supplementary Table 5).

### Analysis of the PedCovid data

We applied the corrected framework F1 to the PedCovid dataset. We checked whether the infection patterns generated by the model matched those in PedCovid. Using 200 simulated datasets with parameters drawn from the posterior distribution, we computed the numbers of infected individuals in each age category and per household for each household size. These were compared to the overall PedCovid statistics. For each household, we used posterior samples of model parameters and augmented data to probabilistically reconstruct transmission trees.

## Results

### PedCovid: strong differences in SARS-CoV-2 infection incidence during the two periods

Household and individual characteristics are given in Table 1. In total, 128 households were recruited during the Alpha period, including 407 contacts (i.e. household members other than the inclusion case), of whom 215 were infected. During the Omicron period, 54 households were recruited, including 180 contacts of whom 127 were infected. Households were of median size 4 (Q1-Q3: 4-5) during both periods. Median ages of inclusion cases were 10 (7-13.25) and 6 (4.25-8) years, during the Alpha and Omicron periods, respectively. During the Omicron period, the median proportion of infected contacts in a household was 93% (50%-100%), with 50% (27/54) of households having all contacts infected. This median proportion was lower during the Alpha period (50%), and only 30% (38/128) had all contacts infected (Supplementary Figure 7).

**Table 1.** Household characteristics and individual characteristics in the PedCovid study for the Alpha and Omicron periods. *Values are reported as median (Q1-Q3).

|  |  | Alpha period |  |  | Omicron period |  |  |
| --- | --- | --- | --- | --- | --- | --- | --- |
| Household characteristics |  |  |  |  |  |  |  |
| Number of households |  | 128 |  |  | 54 |  |  |
| Household size* |  | 4 (4-5) |  |  | 4 (4-5) |  |  |
| Age of inclusion case* |  | 10 (7-13.25) |  |  | 6 (4.25-8) |  |  |
| Number of households declaring control measures | None | 83 (65%) |  |  | 35 (65%) |  |  |
|  | Disinfection | 22 (17%) |  |  | 14 (26%) |  |  |
|  | Isolation | 6 (5%) |  |  | 0 (0%) |  |  |
|  | Both | 17 (13%) |  |  | 5 (9%) |  |  |
| Proportion of infected contacts in a household* |  | 50% (24%-100%) |  |  | 93% (50%-100%) |  |  |
| Individual characteristics |  |  |  |  |  |  |  |
| Number of individuals |  | 535 |  |  | 234 |  |  |
|  |  | Infants (<6yo) | Children (6-11yo) | Teens/adults (>11yo) | Infants (<6yo) | Children (6-11yo) | Teens/adults (>11yo) |
| Inclusion cases | Number of individuals | 21 (16%) | 55 (43%) | 52 (41%) | 24 (44%) | 24 (44%) | 6 (11%) |
|  | Proportion of asymptomatic | 5 (24%) | 16 (29%) | 6 (12%) | 4 (17%) | 5 (21%) | 0 (0%) |
|  | Past exposures | 2 (10%) | 7 (13%) | 10 (19%) | 3 (12%) | 4 (17%) | 6 (100%) |
|  | Including vaccination | 0 (0%) | 1 (2%) | 0 (0%) | 1 (4%) | 1 (4%) | 5 (83%) |
| Household contacts | Number of individuals | 40 (10%) | 69 (17%) | 298 (73%) | 36 (20%) | 25 (14%) | 119 (66%) |
|  | Number of infected individuals | 17 (42%) | 36 (52%) | 162 (54%) | 26 (72%) | 20 (80%) | 81 (68%) |
|  | Proportion of asymptomatic | 5 (29%) | 13 (36%) | 21 (13%) | 6 (23%) | 4 (20%) | 18 (22%) |
|  | Past exposure | 3 (8%) | 10 (14%) | 71 (24%) | 3 (8%) | 7 (28%) | 113 (95%) |
|  | Including vaccination | 0 (0%) | 0 (0%) | 27 (9%) | 0 (0%) | 1 (4%) | 106 (89%) |

Among infected contacts, there was a smaller proportion of infected 0-11 years old children during the Alpha period (25%, 53/215) than the Omicron period (36%, 46/127). The proportion of asymptomatic infections in 0-11 years old infected contacts were 34% (18/53) and 22% (10/46) during the Alpha and Omicron periods, respectively. Only 5% (28/535) of all participants were vaccinated during the Alpha period, 96% (27/28) of them being adults. During the Omicron period, 49% (114/234) of participants were vaccinated, 97% (111/114) being adults.

### Recruitment protocol impacts estimation results

In our simulation study, the recruitment method did not affect the estimation of parameters associated with control measures, past exposure, and the risk of infection from the community (Supplementary Figures 3 and 4). For age-specific infectiousness and susceptibility parameters, results changed with recruitment method and inference framework, as shown in Figure 2 for the Alpha period.

**Figure 2.**
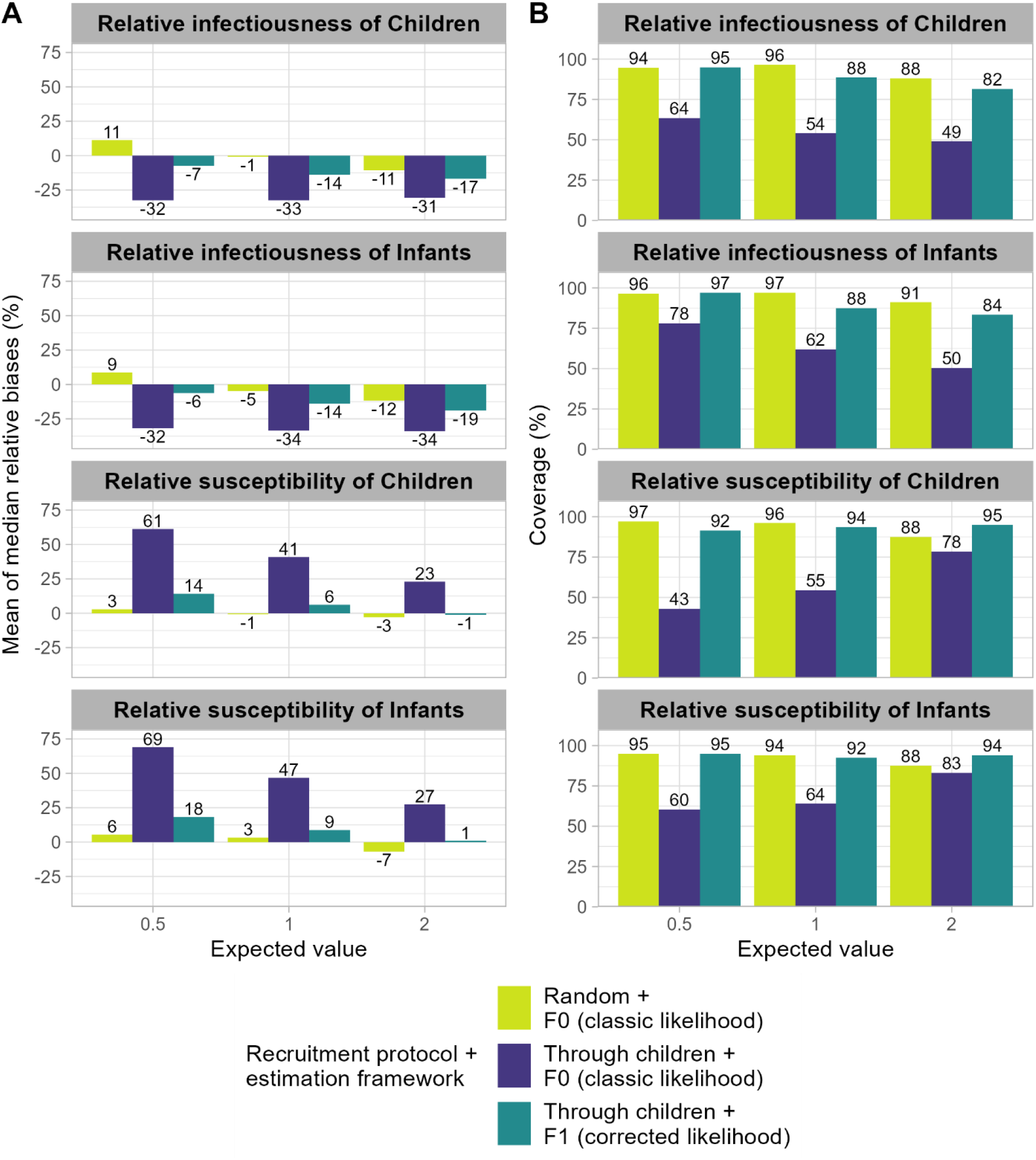
Comparison of the posterior estimates of infectiousness and susceptibility parameters across age groups, for random recruitment and recruitment through children and under the two frameworks (Alpha period). A – Mean of median relative biases across simulations (in percentage) in the three different scenarios. B – Coverage of posterior credible intervals in the three different scenarios, i.e. percentage of the simulations for which the posterior credible interval includes the expected value.

When recruitment was random, no significant bias in infectiousness and susceptibility across age groups was observed overall (Figure 2A), and coverage values reached more than 88% for all parameters and all expected values (Figure 2B). By contrast, for recruitment through positive children, the classic inference framework (F0) led to a systematic underestimation of the infectiousness of children, and overestimation of their susceptibility. For example, biases of 33% to 47% are reported under the null hypothesis (no homogeneous infectiousness and susceptibility across age groups), with coverages of 54% to 64%. The corrected framework (F1) largely corrected for this bias, with biases ranging between 6% and 14% under the null hypothesis for example, and coverage values of 82% to 95% for all parameter values (Figure 2B).

Similar patterns were found for the Omicron period (Supplementary Figure 5).

### Analysis of the PedCovid data

#### Heterogeneous age-specific transmission during the Alpha period but not the Omicron period

Using the inference framework accounting for household recruitment protocol, we estimated global transmission parameters and the effect of age, control measures, and past exposure on susceptibility and infectiousness during the Alpha and Omicron periods in France through the analysis of the PedCovid study (Figure 3).

**Figure 3.**
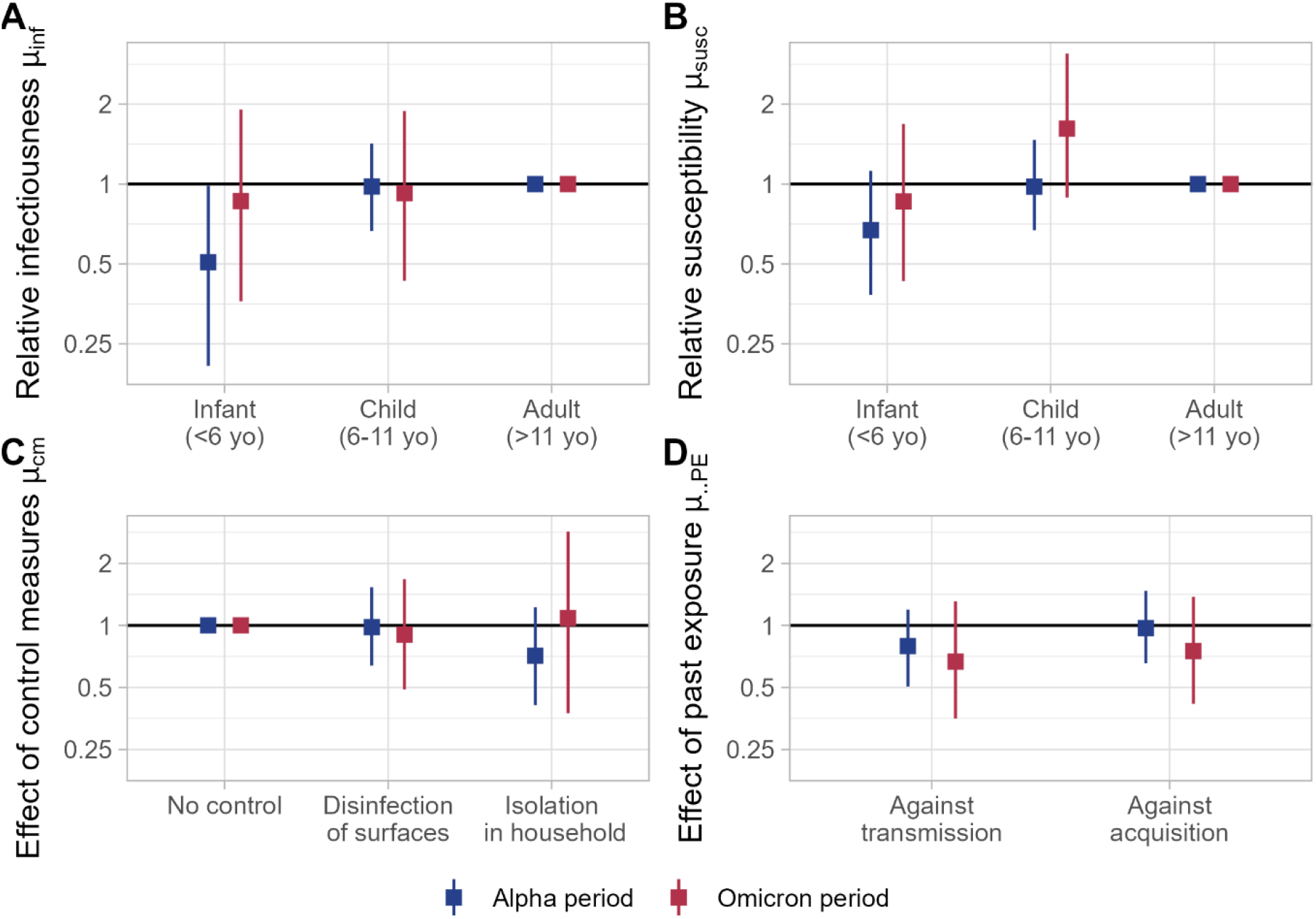
Parameters estimated with the corrected framework F1 from the PedCovid study during the Alpha and Omicron periods. The y-axis is on a logarithmic scale. A – Relative infectiousness across ages μ_infI_ and μ_infc_ (reference: adults). B – Relative susceptibility across ages μ_suscI_ and μ_suscc_ (reference: adults). C – Relative susceptibility across levels of control measures μ_cmDisinf_ and μ_cmIsol_ (reference: no control). D – Effect of past exposure against transmission μ_infPE_ and infection μ_suscPE_ (reference: no past exposure).

#### Age groups

During the Alpha period, the median infectiousness of infants under 6 years old was half that of teenagers and adults (*µ*_*infI*_ =0.51, 95% CrI: 0.21-0.98). The infectiousness of children aged 6-11 years was similar to that of teenagers/adults (*µ*_*infc*_ =0.98, 0.66-1.42). During the Omicron period, we found no age-specific differences in infectiousness (Figure 3A). We estimated no significant age-specific differences in susceptibility for either variant (Figure 3B).

#### Control measures

We showed no impact of declared surface disinfection or isolation of infected cases on transmission in either period (Figure 3C).

#### Past exposures

No significant impact of past exposures on infection or transmission was found (Figure 3D).

The cumulative force of infection of an adult by an infant in a household of size 4 with no control measures and no past exposures reported was 0.27 (0.11-0.50) during the Alpha period, and 0.72 (0.31-1.34) during the Omicron period.

Results were not sensitive to the choice of generation time distribution, duration of test positivity and prior distributions (see Supplementary Figures 12, 13 and 20).

#### Validation and reconstruction of transmission trees

Using the posterior distributions of our parameters, we simulated synthetic outbreaks to assess whether they correctly reproduced the observed data. We show that the model reproduces globally well the numbers of infected individuals in the PedCovid data, both according to age category and household size (Figure 4A and Figure 4B).

**Figure 4.**
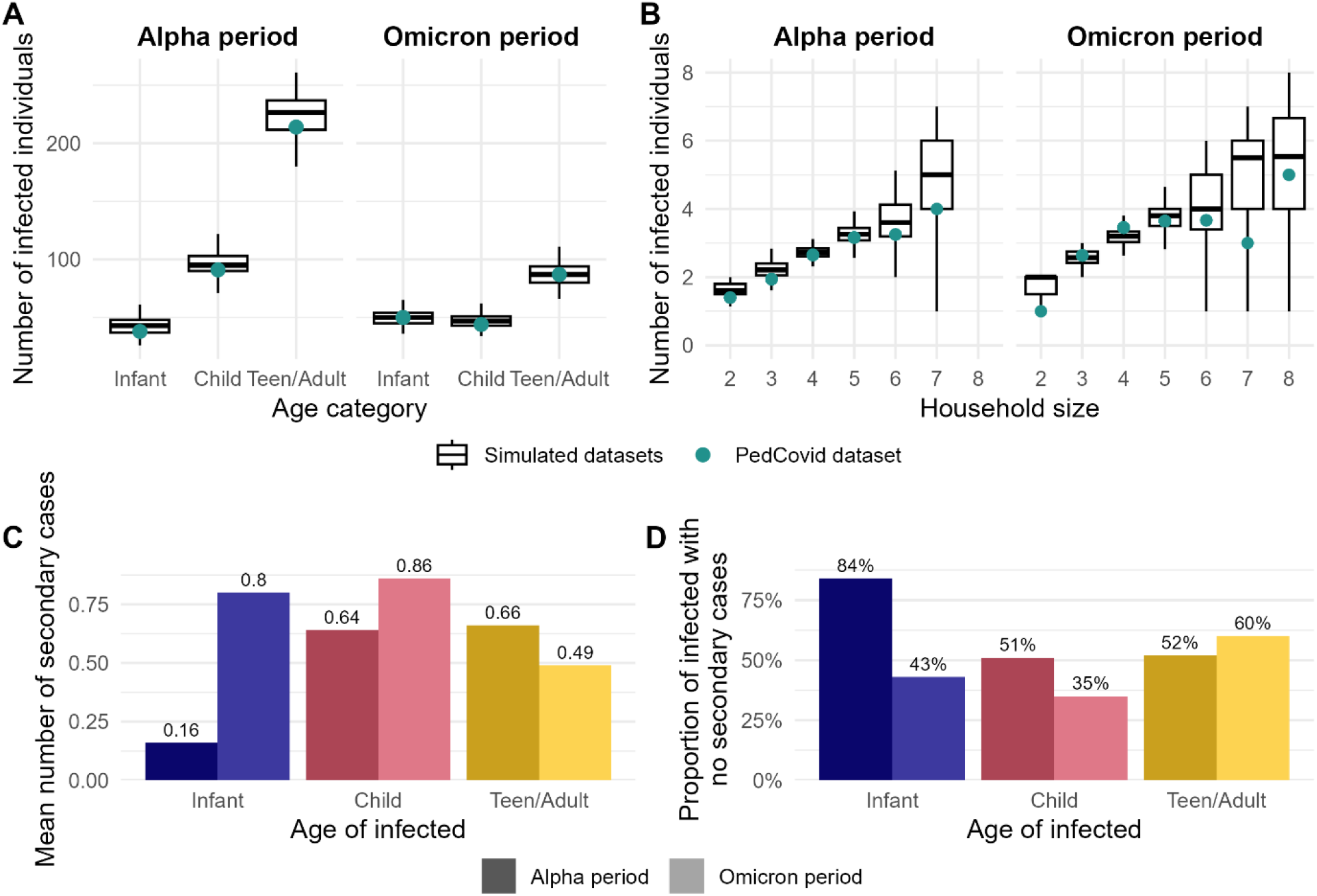
Model adequacy and reconstruction of transmission routes. A – Comparison between simulated and observed numbers of infections by age group. Boxplots show the numbers of infections in datasets simulated using 200 parameter sets sampled from the posterior distribution; green dots indicate the total number of infections in the PedCovid data. B – Comparison between simulated and observed attack rates by household sizes. Boxplots show the attack rate in datasets simulated using 200 parameter sets sampled from the posterior distribution; green dots indicate the mean number of infected individuals over all households of a given size in the PedCovid data. C – Mean number of secondary cases depending on the age group of the infector in the reconstructed transmission chains of the PedCovid data. D – Proportion of infected individuals of each age group that generated no secondary cases in the reconstructed transmission chains of the PedCovid data. Blue: infants (<6 years); pink: children (6-11 years); yellow: teenagers/adults (>11 years).

Finally, we probabilistically reconstructed transmission trees in PedCovid households (example in Supplementary Figure 21). We show that, during the Alpha period, the mean number of secondary cases generated by infants was 0.16 individuals, while it was 0.64 and 0.66 for children and teenagers and adults, respectively (Figure 4C). The proportion of infected infants that generated no secondary cases, either because they did not transmit or because there were no susceptible contacts left, was high during the Alpha period (84%), but reduced to only 43% during the Omicron period (Figure 4D).

## Discussion

In this study, we aimed at analyzing the households of the PedCovid study to assess the age-specific infectiousness and susceptibility of the Alpha and Omicron variants of SARS-CoV-2. This led us to shed light on the estimation bias arising from specific household study designs and the necessity to account for it in models. We thus proposed a correction of the likelihood to reliably estimate transmission parameters. Using our corrected model, we showed that the infectiousness of the Omicron variant was homogeneous across age groups, whereas infants under six years old were less infectious with the Alpha variant than other age groups.

Our simulation study evaluating the bias introduced by the recruitment protocol revealed an underestimation of children’s infectiousness and an overestimation of their susceptibility, when recruiting households through positive children. This bias can be explained by the presence, in any selected household, of at least one infected child, while infected adults may be absent. By contrast, our results demonstrate that recruitment through children does not impact estimates of the effectiveness of control measures or past exposures, as expected since these parameters are not age-dependent in our model.

The bias identified here is similar to that identified by Bell et al^15^. In their study of transmission of human metapneumovirus and seasonal coronaviruses in households recruited through children, they found a negative association between the age of secondary cases and transmission. They attributed this association to selection bias. To our knowledge, only one study accounted for selection bias in their framework: Cousien et al^16^ analyzed Zika virus infections in households recruited based on the detection of symptomatic cases, by accounting for the probability of household detection. Further studies should investigate more systematically the biases introduced by other recruitment strategies (e.g. through adults). Here, we propose a novel approach to correct the likelihood and explicitly account for recruitment by directly computing the theoretical probability of recruitment. Our correction substantially reduced recruitment bias, yielding results close to those obtained under random recruitment. Residual biases of less than 20% remained, however, for extreme parameter values (0.5 and 2). These may be explained by the simplifying assumptions used to compute the likelihood of the recruitment constraint: we assumed that individuals could only be infected by the index case, thereby minimizing the impact of tertiary infections. Nevertheless, validation using posterior distributions showed biases below 12% for all relative parameters (Supplementary Table 5). Other methods, such as simulation-based inference approaches^17^, should also be explored to address selection bias.

Using this new framework, we analyzed the transmission of SARS-CoV-2 in the PedCovid households. We showed a reduced infectiousness of infants compared to adults during the Alpha period, which is consistent with several studies^4,18,19^. We also show no significant difference of susceptibility across age groups. This is consistent with many studies^2,4,18,20^, even though most of them consider broader children age categories (<18 years old). For the Omicron period, we show no significant difference of infectiousness or susceptibility across ages due to large credible intervals that may suggest lack of power. These insights on the role of children in transmission dynamics in households contribute to the body of evidence about age-specific transmission differences of SARS-CoV-2.

The impact of interventions set up at the household level has been poorly investigated over time. Here, we did not evidence any impact of declared surface disinfection or isolation on transmission during either study period. Previous studies reported a significant impact of isolation: Layan et al^6^ found a 80% reduction in infection risk for isolated individuals. Other studies found no impact and suggested that most transmission events occurred shortly after virus importation, meaning that measures were implemented too late^21^. This lack of effect may also suggest that transmission rates were too high, rendering household isolation ineffective, or that isolation was not applied strictly enough. In addition, we made the assumption that control measures applied uniformly to all households declaring measures, and all household members, as soon as infection was detected. Yet, these declarative data may be subject to recall or social desirability biases. In particular, the implementation of control measures may vary across households, periods and age groups (isolation for infants is probably less strict than for older children, or adults). Finally, the number of households applying control measures in our study was limited, restricting statistical power. Studies specifically designed to assess the impact of interventions, with more precise data collection, should be implemented in the future to provide more evidence on that question. More behavioral studies, such as with contact data, would also be helpful to account for behavioral differences in models.

We found no significant protection by past exposure. During the Alpha period, our median estimates for the effectiveness against both infection (3%) and transmission (21%) are lower than those found in the literature. Layan et al^6^ and Prunas et al^22^ showed a 75% and 63% effectiveness against infection, respectively. Oordt-Speets et al^23^ showed that effectiveness against transmission ranged from 39% to 75%. For the Omicron variant, our estimate of effectiveness against transmission (33%) aligns better with estimates from Oordt-Speets et al^23^ (16%-31%), but estimates of effectiveness against infection are again lower than documented^24^. These differences could be explained by the fact that we combined past infections and vaccination, and did not account for waning immunity^25,26^. The timing of previous infections was not known, and for the Omicron period, the delay between vaccination booster and inclusion in PedCovid was longer than 6 months on average. Meggiolaro et al^27^ showed that the reduction against infection is 69% smaller for studies with 3-months follow-up. Our estimates for both periods may therefore depict this waning of immunity.

Many assumptions were made either to keep the model parsimonious or to adapt to the available data. First, we considered contacts to be homogeneous within households. Studies have suggested that household contacts vary by age^28^. Layan et al^29^ showed that ignoring this heterogeneity can lead to an underestimation of children’s susceptibility and infectiousness by up to 20%, which would mitigate the estimated impact of age on Alpha variant transmission. Second, our model does not account for differences in transmission by symptomatic status. Consequently, our age-specific infectiousness parameters represent an average across symptomatic and asymptomatic individuals within each age group. In particular, evidence suggests that asymptomatic individuals tend to be less infectious than symptomatic ones, although estimates are highly heterogeneous^30,31^. A review also reported that the proportion of asymptomatic infections is highest among children, peaking around 13.5 years of age^32^. This likely reduces the estimated infectiousness of children and infants. Third, the PedCovid dataset includes results from both salivary and nasopharyngeal tests. In this analysis, we assumed that both had perfect sensitivity and specificity, which could lead to an underestimation of the number of cases, since we considered that all infections were detected. However, 96% (236/245) of non-infected individuals in our data had at least two tests during the follow-up, confirming their true negativity, so that the risk of false negatives was limited. Finally, the PedCovid study included 128 households during the Alpha period and 54 during the Omicron period, corresponding to 535 and 234 individuals, respectively. These limited sample sizes resulted in substantial uncertainty in some parameter estimates, particularly for the Omicron period analysis.

In conclusion, we highlighted here the importance of accounting for recruitment protocol when estimating infectiousness and susceptibility across ages using mathematical models. We proposed a novel inference framework to analyze data from households recruited through positive children by explicitly accounting for recruitment protocol. We brought out changes in household transmission between variants: lower infectiousness among younger children was observed for Alpha– a pattern no longer seen for Omicron. This study once again illustrates the value of combining household studies with appropriate methodology in providing insights into the transmission of respiratory viruses. Further research exploring possible biases and the statistical power associated with different designs will be crucial for maximizing the potential of these unique tools.

## Data Availability

All data produced in the present study are available upon reasonable request to the authors.

## Acknowledgments

The authors thank the PedCovid working group, including Sylvie Behillil, Naïm Bouazza, Nelly Briand, Agnès Delaunay-Moisan, Flora Donati, Vincent Enouf, Jérémie Guedj, Marianne Leruez-Ville, Lulla Opatowski, Faheemah Padavia, Isabelle Sermet-Gaudelus, Chloé Sturmach, Sylvie van der Werf, and the technical team of the National Reference Center for Respiratory viruses. We also thank Jonas Arruda and Jan Hasenauer for insightful discussions on biases and simulations.

## Funding

S.Ch. acknowledges financial support from Université de Versailles Saint-Quentin-en-Yvelines, the Inception program (Investissement d’Avenir grant ANR-16-CONV-0005), Institut Pasteur, and the HOME project (ANR 20-CE35-0016). S.Ca. acknowledges support by the European Commission under the EU4Health programme 2021-2027, Grant Agreement - Project: 101102733 — DURABLE, the Laboratoire d’Excellence Integrative Biology of Emerging Infectious Diseases program (grant ANR-10-LABX-62-IBEID), and the INCEPTION project (PIA/ANR16-CONV-0005). L.O. acknowledges funding from the MODCOV project from the Fondation de France as part of the alliance framework “Tous unis contre le virus” (#106059), and the French National Research Agency and the “Investissement d’Avenir” program, Laboratoire d’Excellence “Integrative Biology of Emerging Infectious Diseases” (ANR-10-LABX-62-IBEID).

The PedCovid study was funded by a grant from the French Ministry of Health (PHRC) and a grant from the ANR (RA-COVID-19).

## Contributions

S.Ch.: Conceptualization, Data Curation, Methodology, Software, Validation, Writing – original draft, Writing – review & editing. M.L.: Methodology, Software, Writing – review & editing. P-Y.B.: Methodology, Writing – review & editing. J.G.: Methodology, Writing – review & editing. S.W.: Data Curation, Investigation, Writing – review & editing. S.K.: Conceptualization, Supervision, Writing – review & editing. S.Ca.: Conceptualization, Methodology, Supervision, Writing – review & editing. I.S-G.: Conceptualization, Data Curation, Investigation, Writing – review & editing. L.O.: Conceptualization, Methodology, Supervision, Writing – review & editing.

## Conflicts of interest

None

## Notes

### Competing Interest Statement

The authors have declared no competing interest.

### Clinical Trial

NCT04355533

### Author Declarations

This study was approved by the ethics committee of Sud-Mediterranee V (20.04.14.62339).

