## Supplementary for "Estimating age-specific heterogeneity in SARS-CoV-2 transmission from prospective longitudinal studies: the importance of correcting for study design"

*[1]Epidemiology and Modelling of Antimicrobial Evasion (EMAE), Institut Pasteur, Université Paris Cité, Paris, France.*
*[2]CESP, Anti-infective evasion and pharmacoepidemiology team, Université Paris-Saclay, UVSQ, Inserm UMR1018, Montigny-Le-Bretonneux, France.*
*[3]Université Paris Cité et Université Sorbonne Paris Nord, IAME, INSERM, Paris, France.*
*[4]PACRI unit, Institut Pasteur, Conservatoire national des arts et métiers, Paris, France.*
*[5]Modélisation, épidémiologie et surveillance des risques sanitaires (MESuRS), Conservatoire national des arts et métiers, Paris, France.*
*[6]Sorbonne Université, INSERM, Sorbonne Public Health Institute, AP-HP, Hôpital Saint-Antoine, Paris, France.*
*[7]National Reference Center for Respiratory Viruses, Molecular Genetics of RNA Viruses unit, CNRS UMR3569, Institut Pasteur, Université Paris Cité, Paris, France.*
*[8]Equipe de Prévention du Risque Infectieux (EPRI), Assistance Publique - Hôpitaux de Paris (AP-HP), Hôpital Bichat, Paris, France.*
*[9]Université Paris Cité, INSERM U1151, CNRS UMR8253, Institut Necker Enfants Malades, Paris, France; Hôpital Necker Enfants Malades, Centre de Référence Maladies Rares, Paris, France.*
*[10]Mathematical Modelling of Infectious Diseases Unit, Institut Pasteur, Université Paris Cité, INSERM UMR1332, CNRS UMR2000, Paris, France.*
*These authors contributed equally.

### A. Technical details on the estimation framework

We developed an individual-based model of transmission of a respiratory virus in households. The goal was to estimate transmission parameters from the data of the PedCovid longitudinal study; and in particular, to explore the impact of age on transmission and the efficacy of interventions set up by households. Model parameters were estimated using Bayesian Markov chain Monte Carlo methods with data augmentation.

#### A.1. Model parameters

All model parameters and distributions for both SARS-CoV-2 variants are summarized in Supplementary Table 1.

| | Parameter | Unit | Definition | | Value | Source |
|---|---|---|---|---|---|---|
| Estimated | c | /day | Risk of acquisition from community | | Alpha period: estimated<br>Omicron period: estimated | / |
| | β | - | Baseline transmission intensity | | | |
| | δ | - | Dependence of transmission on household size | | | |
| | $\mu_{infC}$ | - | Relative infectiousness | of children (ref: teens/adults) | | |
| | $\mu_{infI}$ | - | | of infants (ref: teens/adults) | | |
| | $\mu_{infPE}$ | - | | according to past exposures (ref: no past exposures) | | |
| | $\mu_{suscC}$ | - | Relative susceptibility | of children (ref: teens/adults) | | |
| | $\mu_{suscI}$ | - | | of infants (ref: teens/adults) | | |
| | $\mu_{suscPE}$ | - | | according to past exposures (ref: no past exposures) | | |
| | $\mu_{cmDisinf}$ | - | Relative susceptibility | according to disinfection (ref: no control measure) | | |
| | $\mu_{cmIsol}$ | - | | according to isolation (ref: no control measure) | | |
| Fixed | f | /day | Generation time distribution | | Alpha period: gamma(shape=2.0, rate=0.44)<br>Omicron period: gamma(shape=3.531, rate=1.098) | Alpha period: An Der Heiden et al.<br>Omicron period: Chen et al. |
| | g | /day | Symptomatic individuals: Incubation period distribution | | Alpha period: gamma(mean=4.42, sd=2.3)<br>Omicron period: gamma(mean=3.09, sd=1.64) | Galmiche et al. |
| | | /day | Asymptomatic individuals: Distribution of the delay from infection to first positive test | | Alpha period: unif(1,14)<br>Omicron period: unif(1,14) | Kojima et al. |
| | $D_{positivity}$ | days | Duration of test positivity | | Alpha period: 14 days<br>Omicron period: 14 days | |

***Supplementary Table 1. List of model parameters for each study period.***

#### A.2. Posterior probability

In the PedCovid study, households are recruited after detection of a positive child (≤18 years old). Usually, studies do not account for the recruitment method in their analysis. Here, we propose a new inference framework (F1) that accounts for this specificity by expressing the posterior probability as $P(\theta, Z|Y, R)$, where $\theta$ is the parameters vector (i.e. $\theta = (c, \beta, \delta, \mu_{infC}, \mu_{infI}, \mu_{infPE}, \mu_{suscC}, \mu_{suscI}, \mu_{suscPE}, \mu_{cmDisinf}, \mu_{cmIsol})$), $Z$ the augmented data, $Y$ the observed data, and $R$ represents the recruitment method of the data. In that case, the posterior probability develops as:

$$P(\theta, \boldsymbol{Z}|\boldsymbol{Y}, R) = \frac{P(\boldsymbol{Y}|\theta, \boldsymbol{Z}, R)P(\theta, \boldsymbol{Z}|R)}{P(\boldsymbol{Y}|R)} = \frac{P(\boldsymbol{Y}|\theta, \boldsymbol{Z}, R)P(\boldsymbol{Z}|\theta, R, \boldsymbol{\tau_1})P(\theta, \boldsymbol{\tau_1}|R)}{P(\boldsymbol{Y}|R)}$$

where $\boldsymbol{\tau_1} = (\tau_{1,H_i})_i$ is the set of augmented infection dates of the index cases (i.e. first infected) of each household $H_i$ (included in $\boldsymbol{Z}$).

$P(\boldsymbol{Y}|R)$ is a constant with respect to $\theta$ and $\boldsymbol{Z}$ and can be omitted in the MCMC.

$P(\theta, \boldsymbol{\tau_1}|R) = P(\theta|R, \boldsymbol{\tau_1})P(\boldsymbol{\tau_1}|R)$ where $P(\theta|R, \boldsymbol{\tau_1})$ represents the prior knowledge on the parameters and is assumed not to depend on the recruitment method or the index case ($\approx P(\theta)$), and $P(\boldsymbol{\tau_1}|R)$ is the prior knowledge on the date of infection of the index case, that we suppose non-informative (uniform) and therefore is a constant with respect to $\theta$ and $\boldsymbol{Z}$ and can be omitted in the MCMC algorithm.

Moreover, $P(\boldsymbol{Y}|\theta, \boldsymbol{Z}, R) \approx P(\boldsymbol{Y}|\boldsymbol{Z})$ since $\boldsymbol{Y}$ only depends on $R$ and $\theta$ through $\boldsymbol{Z}$.

And finally, we have $P(\boldsymbol{Z}|\theta, R, \boldsymbol{\tau_1}) = \frac{P(R|\theta, \boldsymbol{Z}, \boldsymbol{\tau_1})P(\boldsymbol{Z}|\theta, \boldsymbol{\tau_1})}{P(R|\theta, \boldsymbol{\tau_1})}$, with augmented data $\boldsymbol{Z}$ satisfying the recruitment constraint $R$, so that $P(R|\theta, \boldsymbol{Z}, \boldsymbol{\tau_1}) = 1$.

We obtain for corrected framework F1:

$$P(\theta, \boldsymbol{Z}|\boldsymbol{Y}, R) \propto P(\boldsymbol{Y}|\boldsymbol{Z}) \frac{P(\boldsymbol{Z}|\theta, \boldsymbol{\tau_1})}{P(R|\theta, \boldsymbol{\tau_1})} P(\theta)$$

By contrast, for the classic framework (F0), the posterior is simply written, with recruitment omitted:

$$P(\theta, \boldsymbol{Z}|\boldsymbol{Y}) = \frac{P(\boldsymbol{Y}|\theta, \boldsymbol{Z})P(\theta, \boldsymbol{Z})}{P(\boldsymbol{Y})} = \frac{P(\boldsymbol{Y}|\theta, \boldsymbol{Z})P(\boldsymbol{Z}|\theta, \boldsymbol{\tau_1})P(\theta, \boldsymbol{\tau_1})}{P(\boldsymbol{Y})} \propto P(\boldsymbol{Y}|\boldsymbol{Z})P(\boldsymbol{Z}|\theta, \boldsymbol{\tau_1})P(\theta)$$

##### A.3. Likelihood computation

***Likelihood of the observation process***

The observation process $P(\boldsymbol{Y}|\boldsymbol{Z})$ (eq 1) ensures that the augmented data are consistent with the observed data. The continuous date of symptom onset/test positivity $T_i^d$ must be within the observed day $t_i^d$. To account for delays in test positivity, the infection date $\tau_i$ must be later than the day before the last negative test $t_i^{lnt}$, earlier than the day before the first positive test $t_i^{fpt}$, and earlier than $D_{positivity}$ days after the first positive test $t_i^{fpt}$. $D_{positivity}$ was assumed to be 14 days for the analysis. A sensitivity analysis was led to study the impact of this duration on the results.

$P(\boldsymbol{Y}|\boldsymbol{Z}) = \mathbb{1}_{|t^d - T^d|<0.5} \times \mathbb{1}_{(t^{fpt} - D_{positivity} - 0.5) \leq \boldsymbol{\tau} \leq (t^{fpt} - 0.5)} \times \mathbb{1}_{(t^{lnt} - 1.5) \leq \boldsymbol{\tau}}$ (eq 1)

***Likelihood of transmission process***

The likelihood of the transmission process is given in equation 2:

$$\begin{aligned} P(\mathbf{Z}|\theta, \boldsymbol{\tau_1}) &= P(\boldsymbol{T^d}|\boldsymbol{\tau}, \theta)P(\boldsymbol{\tau}|\theta, \boldsymbol{\tau_1}) \quad \text{(eq 2)} \\ &= \prod_{i\ infected} g(T_i^d - \tau_i|s_i) \prod_{\substack{i\ secondary \\ case}} \lambda_i(\tau_i) e^{-\int_{\tau_{1,H_i}}^{\tau_i} \lambda_i(u)\,du} \prod_{\substack{i\ not \\ infected}} e^{-\int_{\tau_{1,H_i}}^{t_{end}} \lambda_i(u)\,du} \end{aligned}$$

The first term is, for all infected individuals $i$, the likelihood of the symptom onset date $T_i^d$ given the infection date $\tau_i$. Distribution $g(\cdot\,|s_i)$ is defined as previously described. The second term is, for all secondary cases $i$, the product of the risk of being infected at time of infection $\tau_i$ and the probability of surviving infection from the first infection time of the household of $i$ $\tau_{1,H_i}$ until time of infection $\tau_i$. The third term is, for all uninfected individuals, the probability of surviving infection from the first infection time of the household of $i$ $\tau_{1,H_i}$ until the end of follow-up $t_{end}$.

***Likelihood of the recruitment protocol***

The previous development of the posterior probability leads to dividing the usual likelihood of the data by the probability of recruitment in each household, given the parameters. Intuitively, this formulation gives less weight in the posterior to households with a high probability of recruitment.

Let us call inclusion individual the household member that leads to the inclusion of the household (infected or not). In a generic household study, a household is recruited upon infection of an inclusion individual. In the PedCovid study, inclusion individuals were infected children: among recruitable households that had an infected inclusion individual, households were recruited only if the inclusion individual was a child. The likelihood of recruitment is therefore computed independently in each household as $P(R|\theta) = P(inclusion\ individual\ is\ underaged \mid inclusion\ individual\ is\ infected)$. Let us note that the observed data do not intervene in this theoretical calculation, so the observed inclusion individual is not known. Let us call $R_{underaged}$ the event "the inclusion individual is underaged", $R_{inf}$ the event "the inclusion individual is infected", and $R_i$ the event "$i$ is the inclusion individual". The likelihood can then be computed as follows:

$$P\left(R\middle|\theta,\boldsymbol{\tau_{1,H}}\right) = P(R_{child} \mid R_{inf},\theta,\boldsymbol{\tau_1})$$

$$= \frac{P(R_{underaged},R_{inf}|\theta,\boldsymbol{\tau_1})}{P\left(R_{inf}|\theta,\boldsymbol{\tau_1}\right)}$$

We then use the law of total probabilities on $(R_i)_i$. In other words, we integrate over all possible inclusion individuals, i.e. all household members.

$$= \frac{\sum_i P\left(R_{underaged},R_{inf}\middle| R_i,\theta,\boldsymbol{\tau_1}\right) P(R_i|\theta,\boldsymbol{\tau_1})}{\sum_j P\left( R_{inf}\middle| R_j,\theta,\boldsymbol{\tau_1}\right) P(R_j|\theta,\boldsymbol{\tau_1})}$$

Knowing that $P\left(R_{underaged}\middle| R_i\right) = 0$ if $i$ is not underaged, we can simplify the upper sum:

$$= \frac{\sum_{i\ underaged} P\left(R_{inf}\middle|R_i,\theta,\boldsymbol{\tau_1}\right) P(R_i|\theta,\boldsymbol{\tau_1})}{\sum_j P\left(R_{inf}\middle|R_j,\theta,\boldsymbol{\tau_1}\right)\ P(R_j|\theta,\boldsymbol{\tau_1})}$$

We consider that each household member has the same probability of being the inclusion individual, so $P(R_i|\theta,\tau_1) = 1/n_{H_i}$, where $n_{H_i}$ is the household size.

$$= \frac{\sum_{i\ underaged} P\left(R_{inf}\middle|R_i,\theta,\boldsymbol{\tau_1}\right)\frac{1}{n_{H_i}}}{\sum_j P\left(R_{inf}\middle|R_j,\theta,\boldsymbol{\tau_1}\right)\frac{1}{n_{H_i}}}$$

We have $P(R_{inf}|R_i, \theta, \boldsymbol{\tau_1}) = 1$ if $i$ is the index case, i.e. $i = index(\tau_1)$.

$$= \frac{\mathbb{1}_{\substack{index(\tau_1)\\ underaged}} + \sum_{\substack{i \neq index(\tau_1),\\ i\ underaged}} P(i\ infected\ before\ time\ of\ inclusion\ t_{inclu}|\theta, \boldsymbol{\tau_1})}{1 + \sum_{j \neq index(\tau_1)} P(j\ infected\ before\ time\ of\ inclusion\ t_{inclu}|\theta, \boldsymbol{\tau_1})}$$

$$= \frac{\mathbb{1}_{index(\tau_1)\ underaged} + \sum_{i \neq index(\tau_1),\ i\ underaged}(1 - e^{-\int_{\tau_1}^{t_{inclu}} \lambda_i(u)\,du})}{1 + \sum_{j \neq index(\tau_1)}(1 - e^{-\int_{\tau_1}^{t_{inclu}} \lambda_j(u)\,du})}$$

Finally, for the simplicity of computation, we make the assumption that individuals are only infected by the index case $index(\tau_1)$ (no tertiary infection).

$$= \frac{\mathbb{1}_{index(\tau_1)\ underaged} + \sum_{i \neq index(\tau_1),\ i\ underaged}(1 - e^{-\int_{\tau_1}^{t_{inclu}}\left(c + h_{index(\tau_1)\to i}(u)\right)du})}{1 + \sum_{j \neq index(\tau_1)}(1 - e^{-\int_{\tau_1}^{t_{inclu}}\left(c + h_{index(\tau_1)\to j}(u)\right)du})}$$

Where $h_{j\to i}(t)$ is the instantaneous risk that infector $j$ infects susceptible $i$ at time $t$ (described in main text eq 2).

##### A.4. Prior distributions

Prior distributions used for each parameter are given in Supplementary Table 2.

| Parameter | Prior distribution | Proposal distribution |
|---|---|---|
| c | uniform(0,0.1) | logNormal |
| β | uniform(0,3) | logNormal |
| δ | uniform(-3,3) | normal |
| $\mu_{infC}$, $\mu_{infI}$, $\mu_{infPE}$, $\mu_{suscC}$, $\mu_{suscI}$, $\mu_{suscPE}$, $\mu_{cmDisinf}$, $\mu_{cmIsol}$ | logNormal(0,1) | logNormal |

***Supplementary Table 2 Prior distributions for each model parameter estimated***

#### B. Technical details on the simulation study

To evaluate the possible bias in parameter estimates due to the household recruitment method, we conducted a simulation study. In this study, outbreaks were generated in silico using our transmission model and the different likelihood-based estimation frameworks were assessed in their ability to recover the different parameters without bias. In practice, a pool of households matching the structure (i.e. socio-demographic and behavioral information) of the households followed up during the PedCovid study was constituted. After importation of the virus by a random initial index case, outbreaks were simulated with a fixed set of parameters. A subset of households was then selected through a recruitment model, and followed up. Two recruitment protocols were evaluated: random selection of households and protocol used in PedCovid, i.e. recruitment through a positive child. More details are provided below on the simulator of outbreaks in a household, and the selection algorithm that models recruitment protocols.

#### B.1. Simulation of household outbreaks

The process of simulation of outbreaks is described in Supplementary Figure 1.

##### B.1.a. Household structures

Outbreaks are simulated in a pool of households with a determined structure containing its size, follow-up duration, measures applied and duration of implementation of measures, and the age and immunity status of each member.

##### B.1.b. Simulation of outbreaks

Outbreaks are simulated in discrete time with daily timesteps.

Let $\theta = (c, \beta, \delta, \mu_{infC}, \mu_{infI}, \mu_{suscC}, \mu_{suscI}, \mu_{infPE}, \mu_{suscPE}, \mu_{cmDisinf}, \mu_{cmIsol})$ be the vector of fixed parameters for a simulation. In each synthetic household, the virus is introduced by a household member randomly sampled (adult or child) and an outbreak is stochastically simulated. During the outbreak simulation, an individual who is infected develops symptoms with probability $1 - p_{asympto}$, where $p_{asympto}$ is fixed (see B.1). If the individual is symptomatic, the date of symptom onset is set from the infection time using the incubation distribution $g$ (Table 1 in the main text).

**Index case**

The index case is chosen randomly in the household and is set to be infected at time $t = 0$.

**Propagation in time**

The outbreak is propagated among household members: on a given day $t$, a susceptible individual can become infected with probability $1 - e^{-\lambda_i(t)}$, where the risk of infection $\lambda_i(t)$ is computed from the parameters $\theta$.

***Symptomatic testing*** If an individual has symptoms onset on day $t$, the testing probability is 1 with a delay between tests and symptoms onset sampled from distribution $d_{sTest}$ (see B.1).

***Test results*** Tests are assumed to be positive from 1 day after infection to 14 days after infection. During this period, we assume 100% sensitivity and specificity of tests, so all infected individuals are detected.

***Household testing*** We assume that when an individual is tested positive at a given time $t$, global testing of the household can occur (that was the case during the COVID-19 pandemic). Each member of their household is therefore tested with a fixed probability $1 - p_{missing}$ and a delay sampled from distribution $d_{hhTest}$ (see B.1).

***Random testing*** If no test is performed in the household at time $t$, each individual has a fixed probability $p_{test}$ (see B.1) of being tested at time $t$ for an independent reason. This process can be the results of contact tracing or precautions, for example.

***Control measures*** During propagation, at the first iteration where any individual has symptoms/positive test, we set the end date of measures using the current iteration as beginning point and duration of implementation of measures.

###### B.1.c. Inclusion and follow-up

The inclusion individual is chosen randomly among individuals with a positive test during the study, so the simulation is started over if no individual has a positive test. The inclusion date is then set from the inclusion case's first positive test date, with a delay sampled from distribution $d_{inclu}$ (see B.1). After inclusion, the household can be followed up according to the desired design and test results can be added.

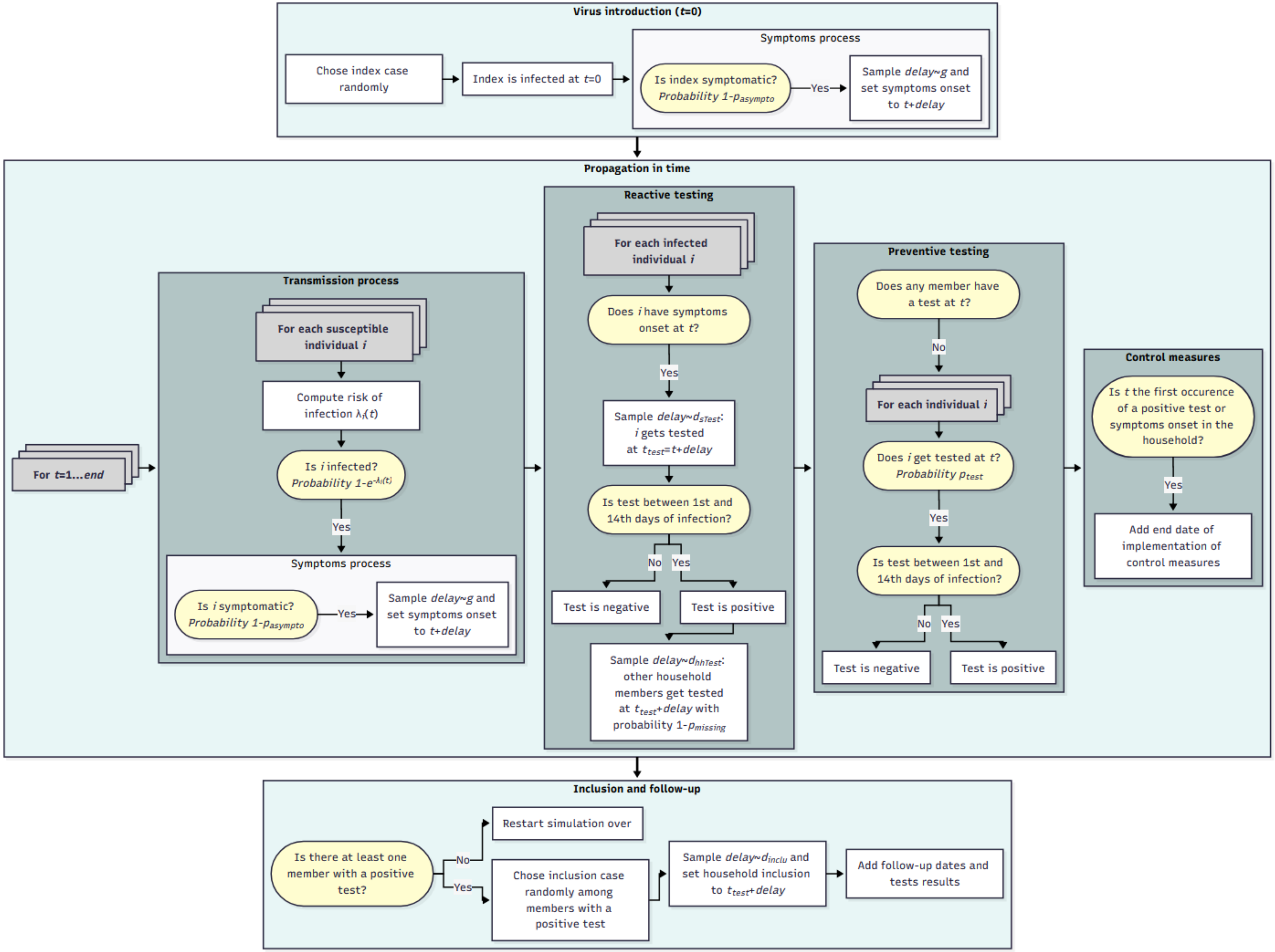

*Supplementary Figure 1. Flowchart of simulation of an outbreak in a household, from virus introduction to inclusion in a study.*

#### B.2. Application to PedCovid

In our analysis, the pool of household structures was created by generating duplicates of each household followed up in the PedCovid study. Each duplicate has the same structure as the original household, with follow-up duration defined as the delay from symptoms/first positive test to last visit, and duration of implementation of measures defined as the shortest delay from first symptoms/positive test of the household to end of measures declared.

All fixed probabilities and distributions used for the simulation are given in Supplementary Table 3.

The distribution of the delay from symptoms onset to test was obtained by fitting a geometric distribution to the observed data on positive delays between date of symptoms onset and date of test before inclusion. The distribution of the delay from the initial positive test to household testing was obtained by fitting a geometric distribution to the observed data on positive delays between date of first positive test before inclusion in a household and dates of tests before inclusion of other household members. Finally, the distribution of the delay from first positive test to inclusion was obtained by fitting a Poisson distribution to the observed data on positive delays between date of first positive test and date of inclusion for inclusion cases. All distributions were fitted to the data using maximum likelihood (R function fitdist from the fitdistrplus package).

Like in the PedCovid study, households were followed up for 45 days, with visits at days 3, 7, 15, and 45. Visits were conducted only if their date fell before their end of follow-up, defined as the date of first detection plus the household-specific follow-up duration specified in the household structure.

| Parameter | Definition | Value | | Source |
|---|---|---|---|---|
| | | Alpha period | Omicron period | |
| $p_{asympto}$ | Proportion of asymptomatic individuals | 0.4 | 0.3 | Fixed |
| $p_{sTest}$ | Probability of symptomatic testing | 1 | 1 | Fixed |
| $d_{sTest}$ | Distribution of the delay from symptoms onset to test (/day) | geometric(proba=0.33) | geometric(proba=0.333) | Fitted from data |
| $p_{missing}$ | Individual probability of missing a household testing | Depends on household size:<br>hsize=2: 0<br>hsize=3: 0.098<br>hsize=4: 0.072<br>hsize=5: 0.135<br>hsize=6: 0.104<br>hsize=7: 0.714 | Depends on household size:<br>hsize=2: 0<br>hsize=3: 0.394<br>hsize=4: 0.239<br>hsize=5: 0.171<br>hsize=6: 0.167<br>hsize=7: 0.143<br>hsize=8: 0.125 | Fitted from data |
| $d_{hhTest}$ | Distribution of the delay from initial positive test to household testing (/day) | geometric(proba=0.481) | geometric(proba=0.463) | Fitted from data |
| $p_{test}$ | Probability of preventive testing | 1/21 | 1/14 | Fixed |
| $d_{inclu}$ | Distribution of the delay from detection to inclusion (/day) | poisson(lambda=4.781) | poisson(lambda=2.667) | Fitted from data |
| $D_{positivity}$ | Duration of test positivity | 14 days | 14 days | Fixed |

***Supplementary Table 3. Value and source of fixed parameters for the simulation study***

#### B.3. Selection of households

From all simulated households, two datasets were generated: one selecting households based on a recruitment through children and one selecting households assuming a random recruitment (see Supplementary Figure 2). For each dataset, 128 households were recruited for the alpha period and 54 households were recruited for the omicron period.

***Recruitment through children.*** A household was included only if the inclusion case was a child.

***Random recruitment.*** Households were included regardless of the characteristics of the inclusion case.

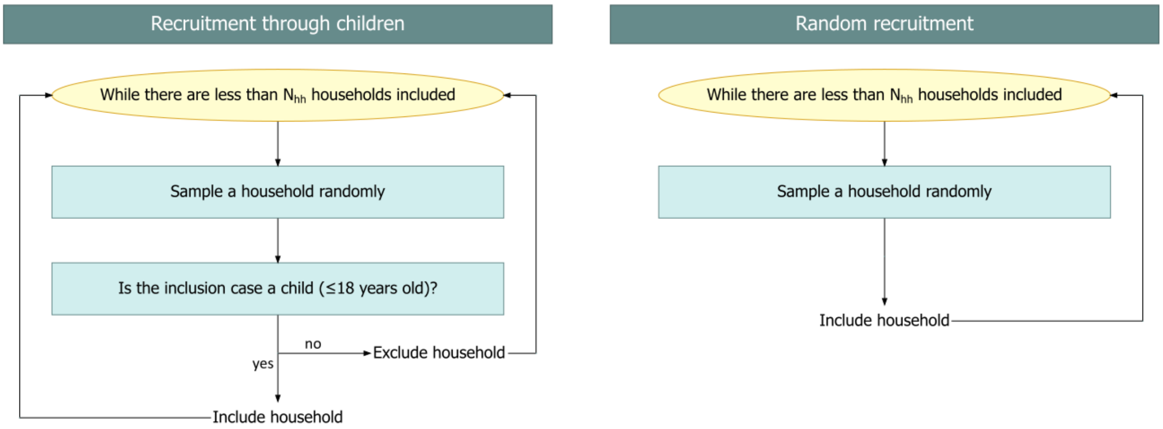

***Supplementary Figure 2. Flowchart of selection of households in the simulation study, for the recruitment through children (left) and the random recruitment (right).*** *$N_{hh}$ is set to 128 for the alpha period and 54 for the omicron period.*

#### B.4. Estimation results

For one fixed set of parameters, we generated 200 independent datasets for each recruitment scenario. For each dataset, we estimated the parameters using the two statistical frameworks: with (F1) and without (F0) accounting for recruitment protocol. Obtained values were compared to the ones used to simulate the data.

##### B.4.a. Evaluation for the null hypothesis of no heterogeneity in transmission.

The first set of parameters assessed assumed no effect of age, symptoms, control measures or immunity on the transmission, i.e. $\mu_{infC} = \mu_{infI} = \mu_{suscC} = \mu_{suscI} = \mu_{infPE} = \mu_{suscPE} = \mu_{cmDisinf} = \mu_{cmIsol} = 1$. Datasets for each recruitment scenario were simulated using the baseline values in Supplementary Table 4.

| Parameter | Baseline values | | Explored values |
|---|---|---|---|
| | Alpha period | Omicron period | |
| c | 0.001 | 0.002 | 0.001 - 0.002 - 0.003 - 0.004 - 0.005 |
| β | 0.429 | 0.551 | 0.2 - 0.5 - 0.8 |
| δ | 1 | 1.372 | 0.4 - 0.7 - 1 - 1.3 - 1.6 |
| $\mu_{infC}$, $\mu_{infI}$, $\mu_{suscC}$, $\mu_{suscI}$ | 1 | 1 | 0.5 - 1 - 2 |
| $\mu_{infPE}$, $\mu_{suscPE}$, $\mu_{cmDisinf}$, $\mu_{cmIsol}$ | | | 0.4 - 0.7 - 1 - 1.3 |

***Supplementary Table 4. Parameter values used in the simulation study. For each parameter, the baseline value and the range over which it is varied are both given.***

Estimation results for each statistical framework are given in Supplementary Figure 3 and Supplementary Figure 4 for the alpha and the omicron variant, respectively.

For the Omicron period, we have biases of 19% to 30%, with coverage values as low as 85%. The framework proposed here, which accounts for the recruitment in the likelihood (F1), largely corrects this bias (Supplementary Figures 3C and 4C). For the Omicron period, the biases on the estimates of the infectiousness parameters are reduced to less than 19% with a coverage of more than 93%, and the biases on the estimates of the susceptibility parameters are reduced to 3%, with a coverage of more than 96%.

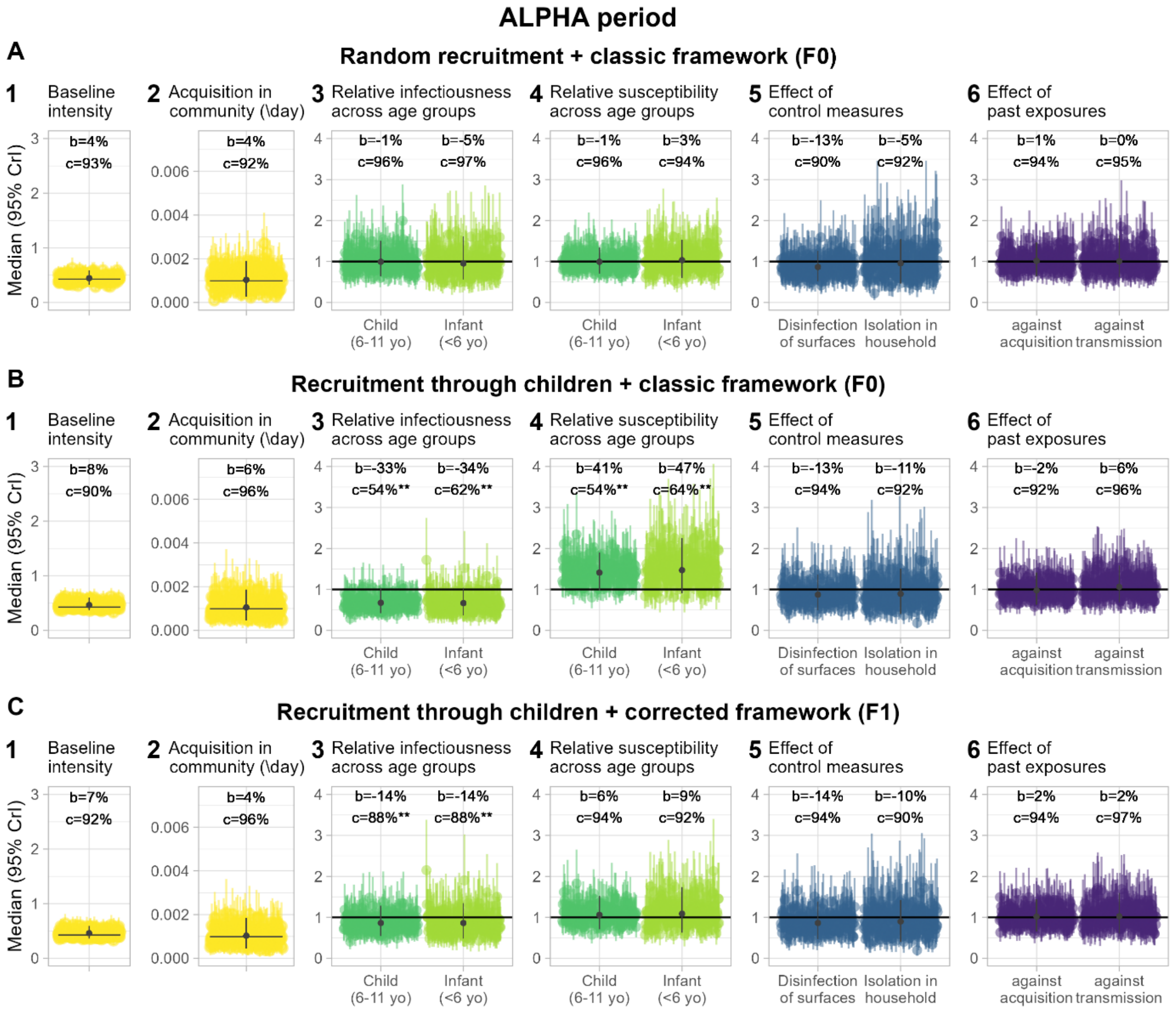

***Supplementary Figure 3. Simulation study results for the Alpha period under the null hypothesis (baseline values of parameters, no difference in infectiousness or susceptibility across individuals). A – Estimations using F0 (framework not accounting for recruitment method) for a dataset generated with a random recruitment scenario. B – Estimations using F0 for a dataset simulated with a recruitment through children. C – Estimations using F1 (framework accounting for recruitment method) for a dataset simulated with a recruitment through children.*** *Color points and intervals indicate the median and 2.5$^{th}$-97.5$^{th}$ percentiles posterior value of the 200 independent simulations. Black points and intervals indicate the distribution (mean and 2.5$^{th}$-97.5$^{th}$ percentiles) of medians over simulations. b=median relative bias in %. c=coverage in %. **: less than 90% of coverage. Yellow: transmission parameters; green: age categories; blue: control measures; purple: past exposure.*

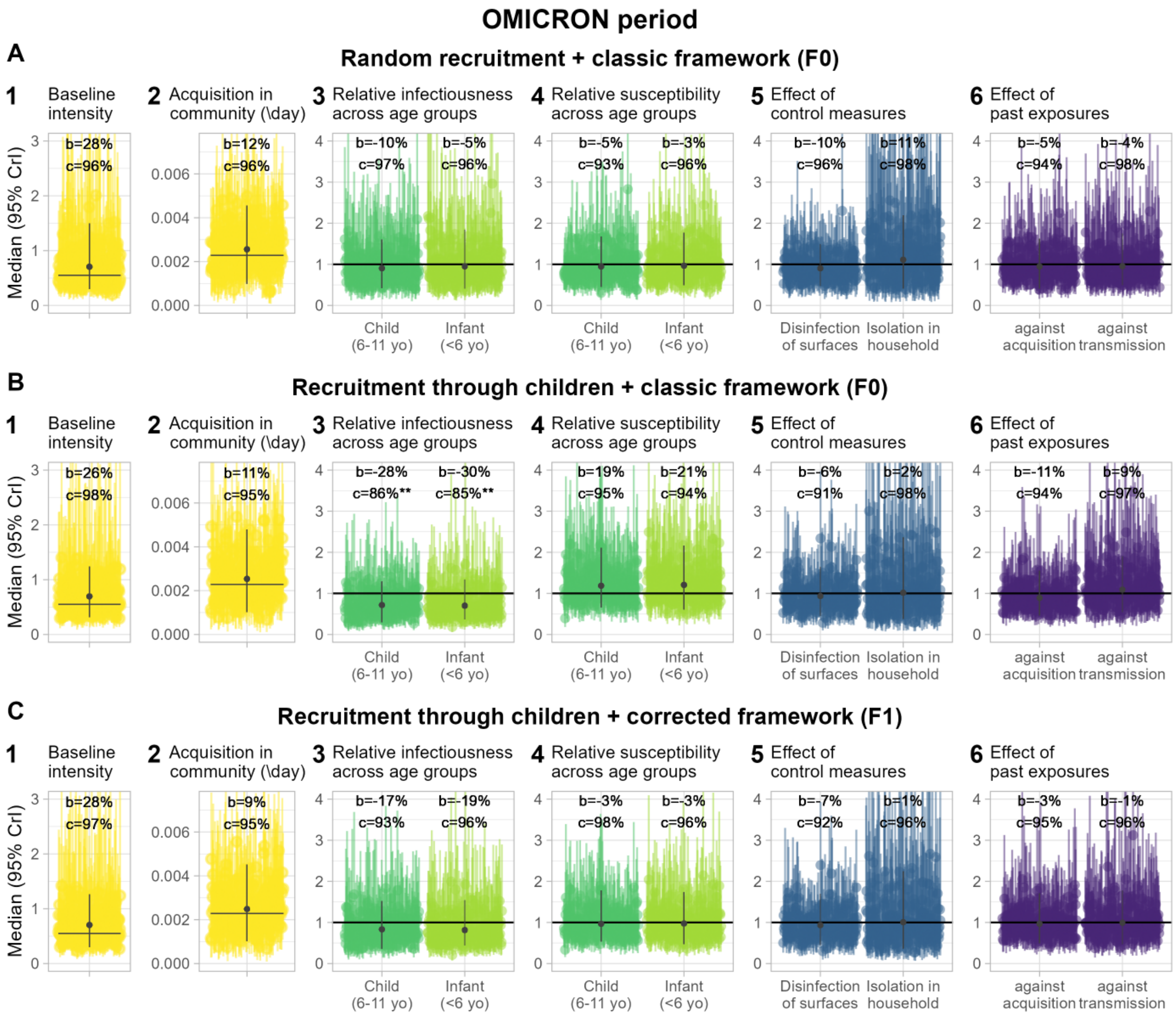

***Supplementary Figure 4. Simulation study results for the Omicron period under the null hypothesis (baseline values of parameters, no difference in infectiousness or susceptibility across individuals). A – Estimations using F0 (framework not accounting for recruitment method) for a dataset generated with a random recruitment scenario. B – Estimations using F0 for a dataset simulated with a recruitment through children. C – Estimations using F1 (framework accounting for recruitment method) for a dataset simulated with a recruitment through children.*** *Color points and intervals indicate the median and 2.5$^{th}$-97.5$^{th}$ percentiles posterior value of the 200 independent simulations. Black points and intervals indicate the distribution (mean and 2.5$^{th}$-97.5$^{th}$ percentiles) of medians over simulations. b=median relative bias in %. c=coverage in %. **: less than 90% of coverage. Yellow: transmission parameters; green: age categories; blue: control measures; purple: past exposure.*

##### B.4.b. Evaluation in case of heterogeneous transmission by age

Other parameter scenarios are defined with baseline values for all parameters except one which is varied according to the range of variations described in Supplementary Table 4. We give here the distribution of median relative biases and coverages comparing framework F0 for random recruitment with the use of both frameworks for recruitment through children, for the Omicron period.

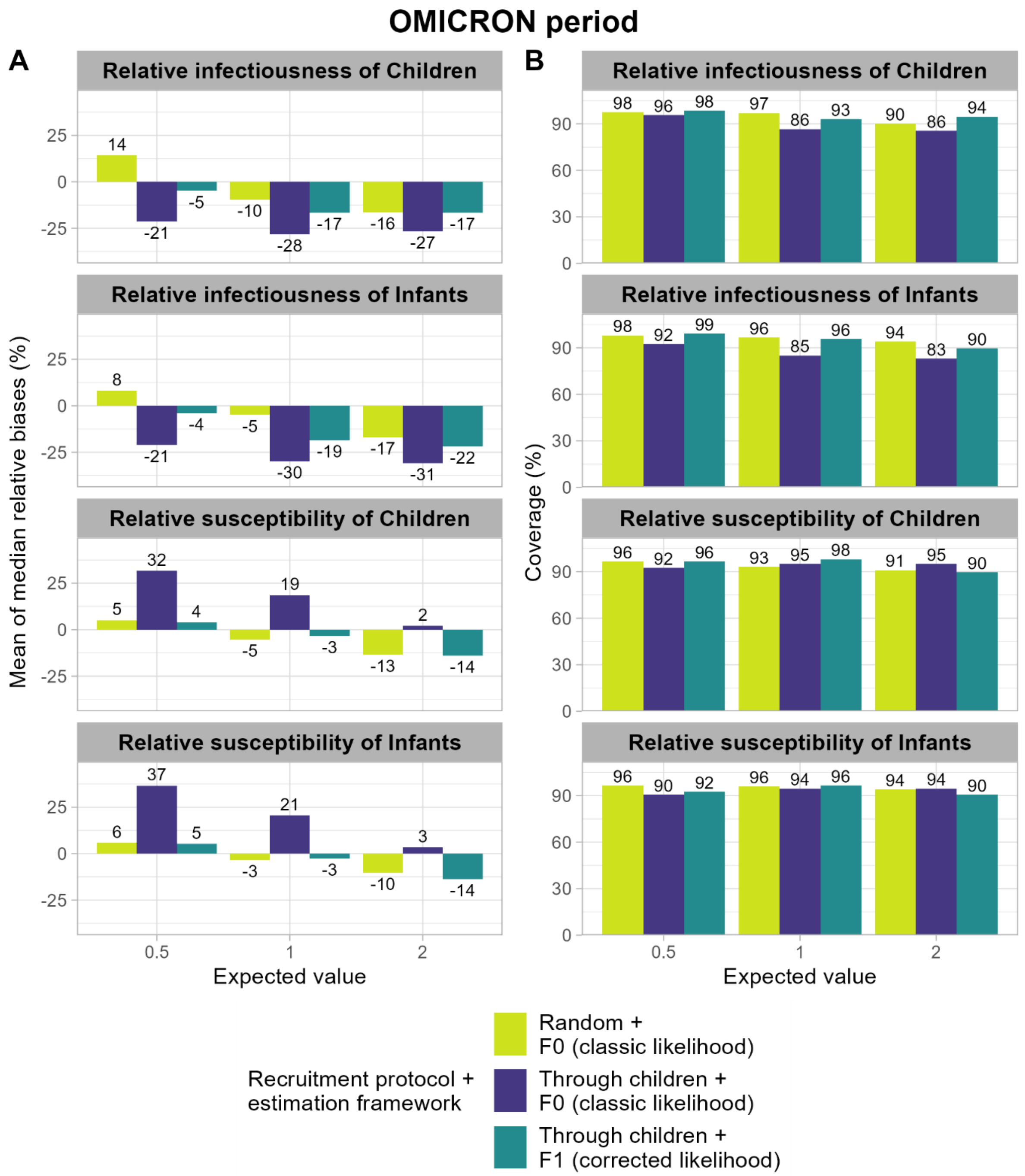

***Supplementary Figure 5. Comparison of the posterior estimates of infectiousness and susceptibility parameters across age groups, for random recruitment and recruitment through children and under the two frameworks (Omicron period). A – Mean of median relative biases across simulations (in percentage) in the three different scenarios. B – Coverage of posterior credible intervals in the three different scenarios, i.e. percentage of the simulations for which the posterior credible interval includes the expected value.***

##### B.4.c. Multivariate validation using posterior distribution from PedCovid data

We finally validated the estimates from the PedCovid dataset by performing a multivariate analysis. We randomly sampled 200 parameter sets in the posterior distributions obtained from the PedCovid dataset. We then simulated the 200 independent associated datasets, assuming a recruitment through positive children, and reanalyzed them through framework F1 to evaluate the accuracy of the estimates. We show here the results in the form of relative bias, see Supplementary Table 5. The bias is close to 0 for all parameters. For the Omicron period, there is a slight over-estimation of the baseline transmission intensity.

| Parameter | **Alpha period** | **Omicron period** |
|---|---|---|
| Community acquisition | -0.05 [-0.7; 0.84] (7%) | 0.17 [-0.59; 1.4] (9%) |
| Baseline transmission intensity | 0.08 [-0.16; 0.33] (7.5%) | 0.31 [-0.49; 1.57] (7.5%) |
| Dependence on hh size | 0.11 [-0.67; 1.32] (5.5%) | 0.06 [-0.63; 0.87] (5.5%) |
| Relative infectiousness across age groups | | |
| Child (6-11 yo) | -0.11 [-0.44; 0.24] (10.5%) | -0.06 [-0.62; 0.84] (7.5%) |
| Infant (<6 yo) | 0.03 [-0.45; 0.86] (3%) | -0.08 [-0.61; 1.17] (7%) |
| Relative susceptibility across age groups | | |
| Child (6-11 yo) | 0.03 [-0.27; 0.42] (4%) | -0.11 [-0.57; 0.72] (9.5%) |
| Infant (<6 yo) | 0.08 [-0.4; 0.88] (5%) | -0.01 [-0.52; 0.8] (3.5%) |
| Effect of control measures | | |
| Disinfection of surfaces | -0.12 [-0.46; 0.42] (8.5%) | -0.08 [-0.53; 0.59] (7.5%) |
| Isolation in household | 0.04 [-0.53; 0.93] (6.5%) | 0.04 [-0.68; 1.37] (3.5%) |
| Effect of past exposure | | |
| against transmission | 0.08 [-0.39; 0.66] (3.5%) | 0.08 [-0.49; 1.13] (5%) |
| against acquisition | -0.04 [-0.33; 0.43] (6%) | -0.01 [-0.46; 0.58] (2.5%) |

***Supplementary Table 5. Median relative bias and rate of type I error for 200 parameter sets sampled from the posterior from the PedCovid data analysis.*** *The table provides the mean and 2.5$^{th}$-97.5$^{th}$ percentiles of the median relative bias, and the percentage of simulations for which the posterior credible interval does not include the real value. The datasets used were simulated using 200 parameter sets sampled from the posterior from the PedCovid data analysis, and a recruitment through children. The estimation framework used is the corrected framework (F1).*

#### C. Analysis of the PedCovid study

##### C.1. Data description

The distribution of ages in inclusion cases and in family members are given in Supplementary Figure 6 for both periods. The distribution of attack rates, defined as the total proportion of individuals infected throughout the study in a household, is given in Supplementary Figure 7 for both periods.

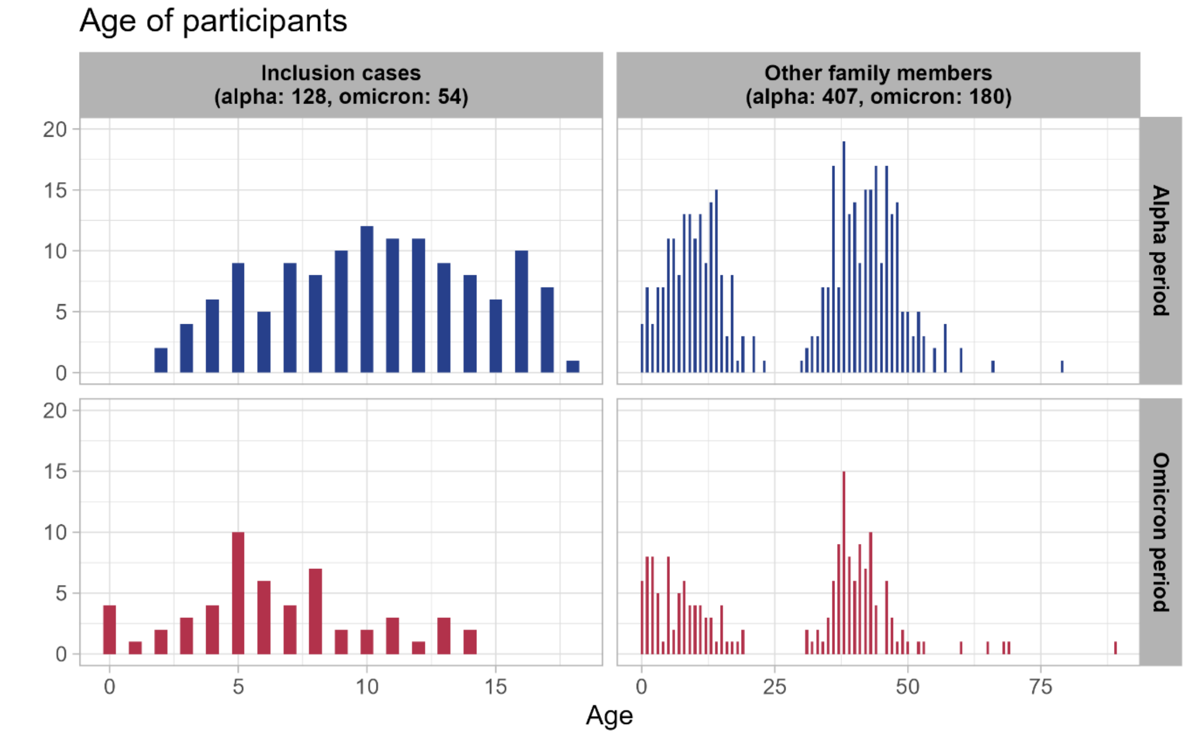

*Supplementary Figure 6. Age distribution of participants for both periods (alpha in blue, omicron in red), for inclusion cases and other family members.*

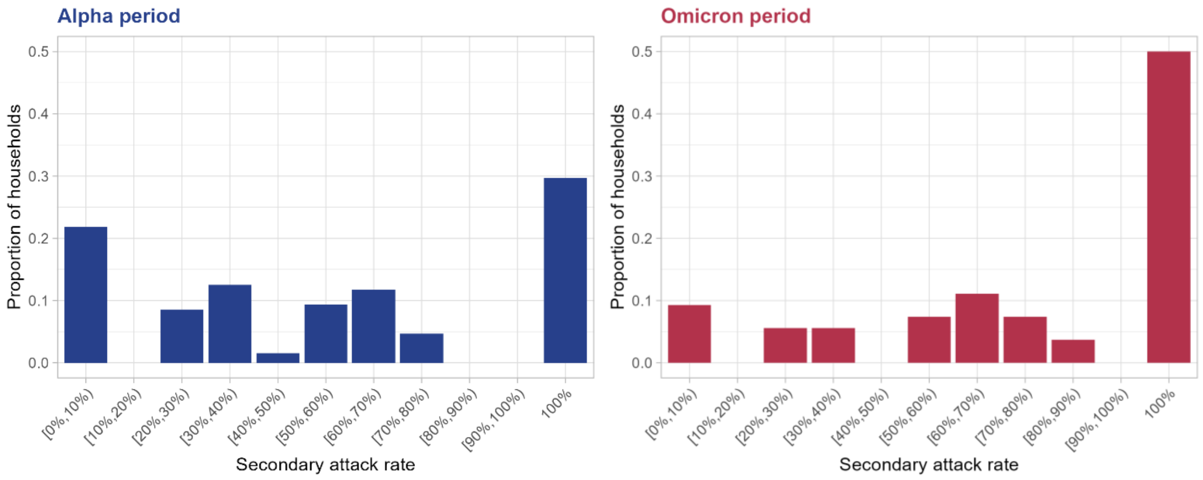

*Supplementary Figure 7. Distribution of secondary attack rates (proportion of household contacts that have been infected during the study) for both periods (alpha in blue, omicron in red).*

An example of a typical trajectory in a household is given in Supplementary Figure 8. For each household member, the results of the different tests, done during visits or outside visits, are shown, as well as potential onset of symptoms, and period of implementation of control measures.

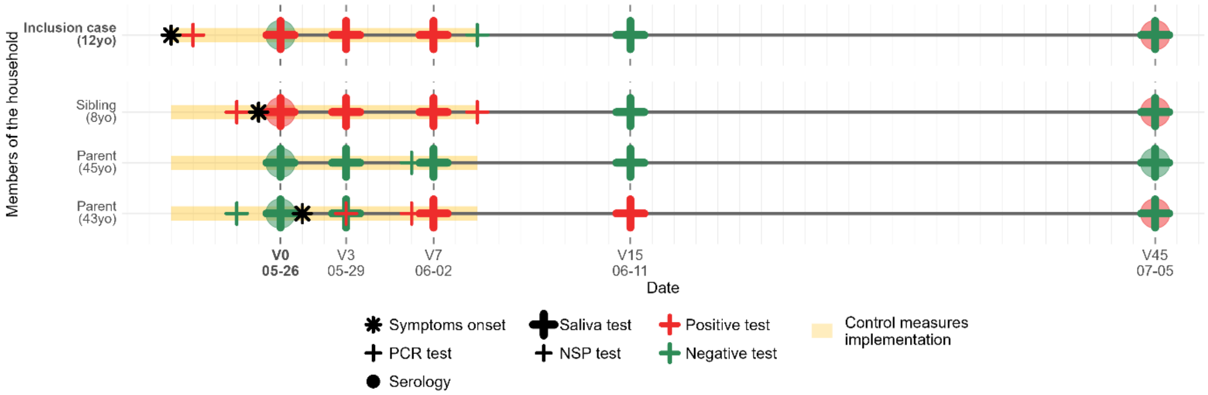

***Supplementary Figure 8. Example of a typical trajectory in a household. Dates and results of PCR and serological tests are given, as well as symptoms onset and period of implementation of control measures.***

#### C.2. Hypotheses about data

Several assumptions were made to analyze the PedCovid data. First, individuals with at least one positive PCR test, either on a nasopharyngeal or salivary sample, throughout the study period were considered as confirmed cases. Asymptomatic cases were defined as confirmed cases who reported no symptoms during the study period, or confirmed cases who reported symptoms more than 10 days before their first positive test. Individuals were considered immunized if they had been vaccinated with a first dose at least 10 days and less than 3 months before inclusion, if they had been vaccinated with a second dose at least 10 days before inclusion, if they had a previous infection more than 1 month and less than 6 months before, or if they had a positive serology at inclusion. We assumed that control measures applied to all individuals of the household, from the first symptoms onset or positive test in the household until the earliest end of measures declared to the nurse.

#### C.3. Results of model zero

Model zero is the simplest model, in which only $c$, $\beta$ and $\delta$ are estimated, and we consider no difference across ages, symptoms, control measures or immunity. The estimates for $c$, $\beta$, $\delta$ for model zero are provided in Supplementary Table 6.

| Parameter | Alpha period | Omicron period |
|---|---|---|
| c | 0.001 [4e-04-0.0018] | 0.002 [6e-04-0.0049] |
| beta | 0.43 [0.37-0.49] | 0.55 [0.45-0.67] |
| delta | 1 [0.34-1.67] | 1.37 [0.67-2.02] |

***Supplementary Table 6. Posterior estimates (median and 95% Credible Interval) from PedCovid data using model zero.***

#### C.4. Estimates for global transmission parameters

The estimated transmission intensity $\beta$, i.e. the cumulative hazard of transmission from an adult to another adult over the full duration of infection, in the baseline case of no control measures nor past exposure, was 0.53 (95% Credible Interval: 0.40-0.67) during the Alpha period, and 0.80 (0.37-1.7) during the Omicron period (Supplementary Figure 9A). The rate of transmission from the community, $c$, was 0.0010 day$^{-1}$ (0.0005-0.0018) during the Alpha period and 0.0024 day$^{-1}$ (0.0007-0.0057) during the Omicron period (Supplementary Figure 9B).

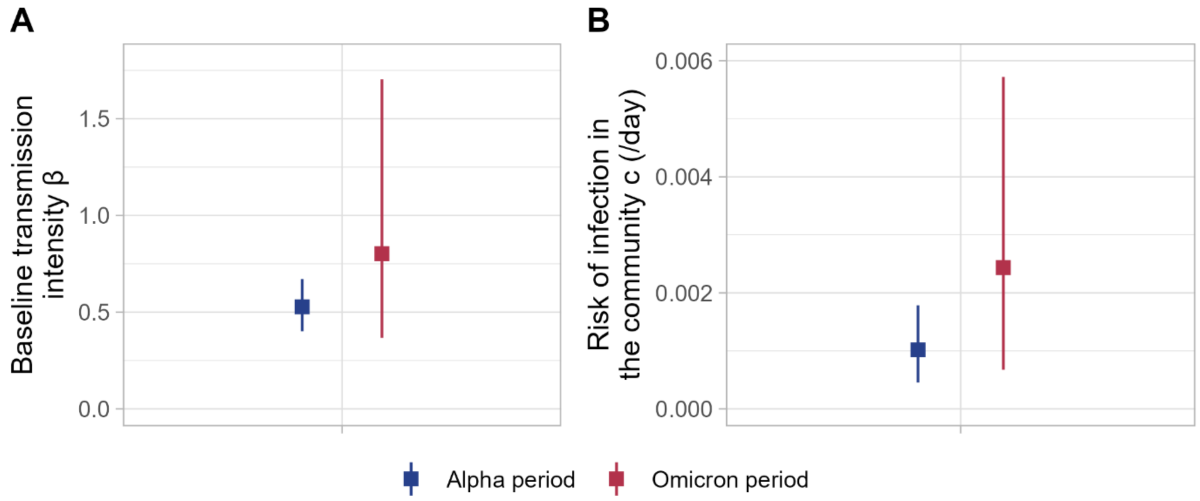

***Supplementary Figure 9. Parameters estimated with the corrected framework F1 from the PedCovid study during the Alpha and Omicron periods. A – Baseline transmission intensity $\beta$. B – Risk of infection in the community $c$ (/day).***

#### C.5. Results for all scenarios

Given the results of our simulation study, we chose to present the results using the model accounting for recruitment protocol (F1). We give the results for the two frameworks F0 and F1 in Supplementary Figure 10. The main difference appears for the relative susceptibility across ages.

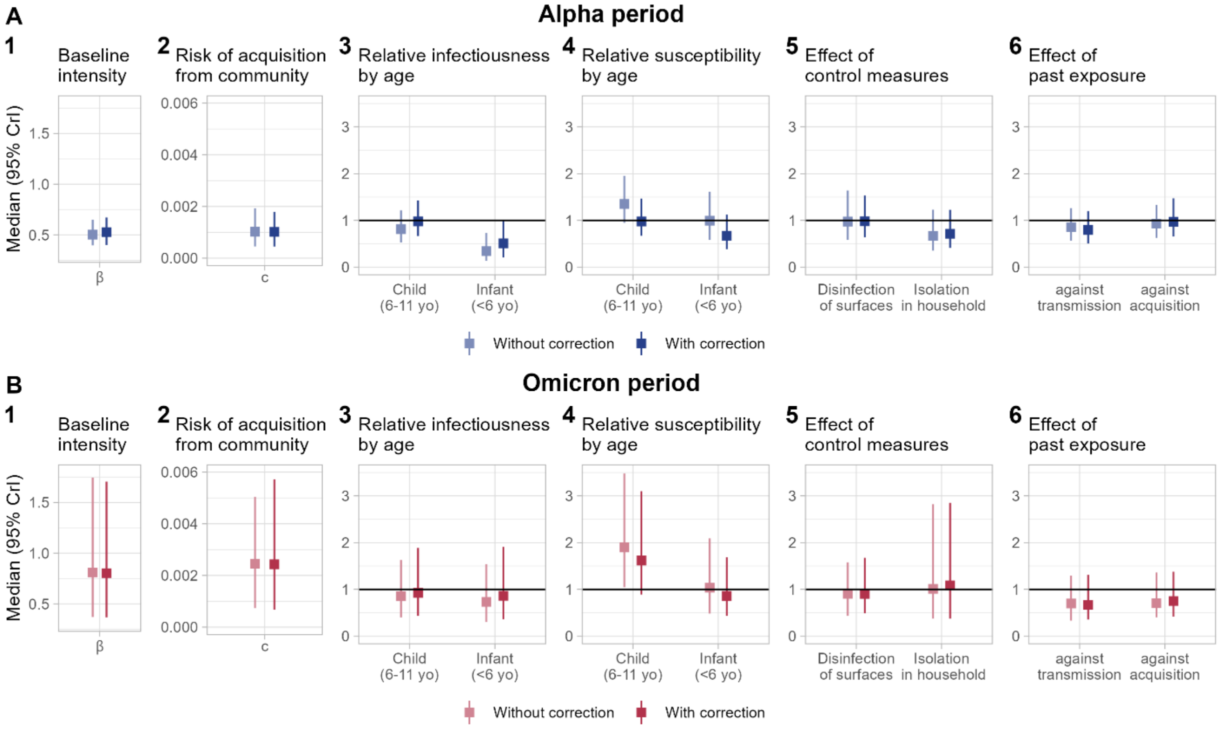

***Supplementary Figure 10. Estimation results without and with correction.*** *Stars indicate parameters that were shown to be biased with more than 10% of false positives by the simulation study of the null hypothesis.*

#### C.6. Sensitivity analysis on the generation time distribution

In the main analysis, we used generation time distributions from the literature. We explored as a sensitivity analysis the impact of shorter or longer generation times, by varying the scale of the gamma distribution. Resulting distributions are given in Supplementary Table 7 and Supplementary Figure 11.

| | Alpha period | | | Omicron period | | |
|---|---|---|---|---|---|---|
| | Distribution | Mean | SD | Distribution | Mean | SD |
| **Baseline** | Gamma(2,1/0.44) | 4.5 | 10.3 | Gamma(3.531,1/1.098) | 3.2 | 2.9 |
| **Shorter** | Gamma(2,1.5) | 3.0 | 4.5 | Gamma(3.531,0.6) | 2.1 | 1.3 |
| **Longer** | Gamma(2,3) | 6.0 | 18.0 | Gamma(3.531,1.2) | 4.2 | 5.1 |

***Supplementary Table 7. Distributions of the generation time for the sensitivity analysis for each period.***

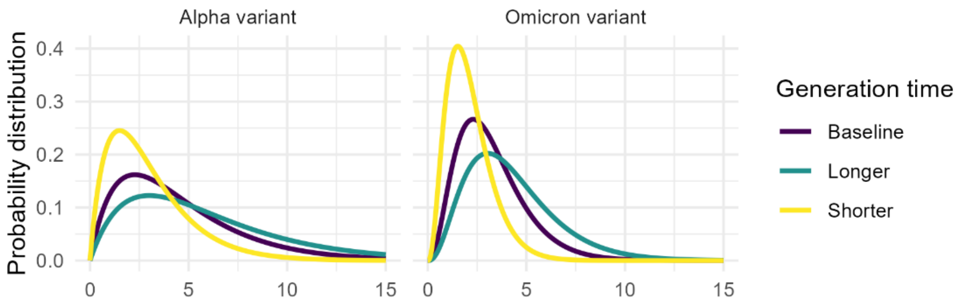

***Supplementary Figure 11. Sensitivity analysis on the distributions of the generation time. Three distributions were explored with baseline, lower and higher values of the scale parameter of the gamma distribution.***

Estimates for each parameter and each distribution are given in Supplementary Figure 12. Results are not sensitive to the distribution of the generation time.

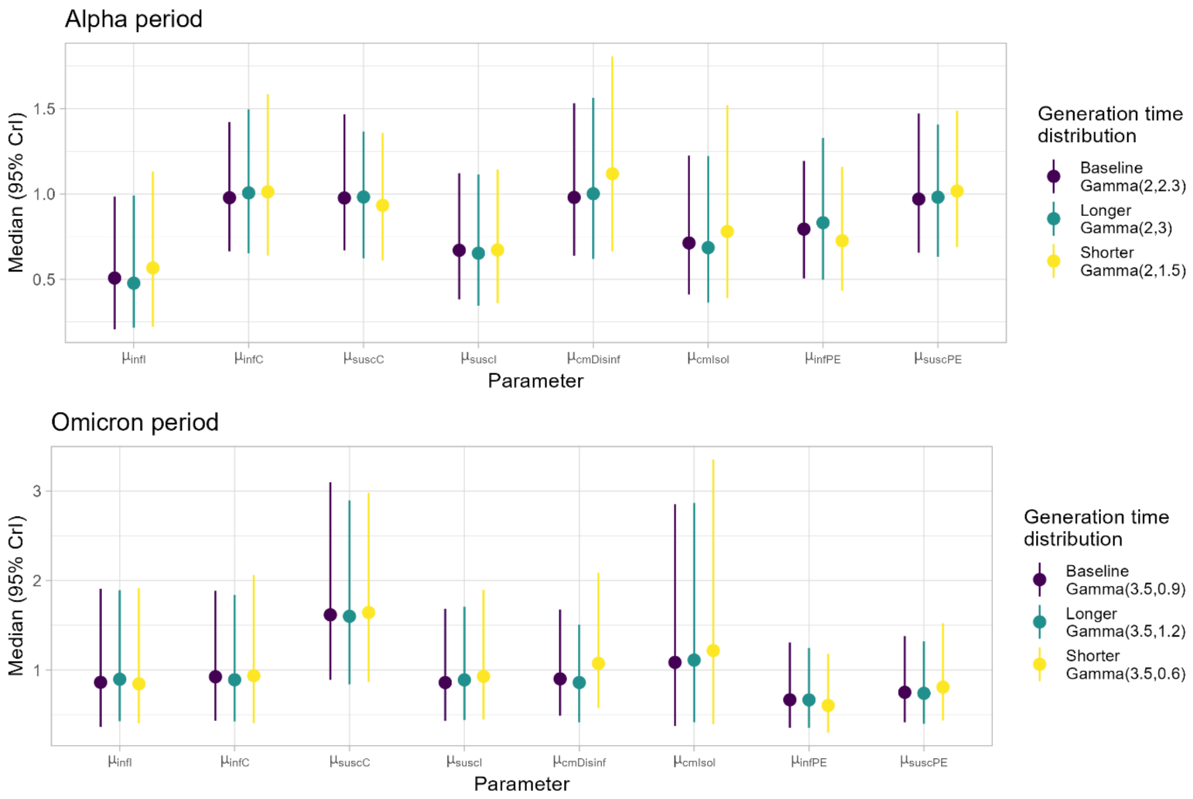

***Supplementary Figure 12. Results of the sensitivity analysis on generation time: median estimates of infectiousness and susceptibility parameters for 3 gamma distributions of varied scale.***

#### C.7. Sensitivity analysis on test positivity duration

In the main analysis, we used a positivity duration of tests of 14 days. We evaluated the impact of this choice of test positivity duration in a sensitivity analysis. We explored durations of 10 and 7 days. Results are given in Supplementary Figure 13. Results are not sensitive to the choice of test positivity duration.

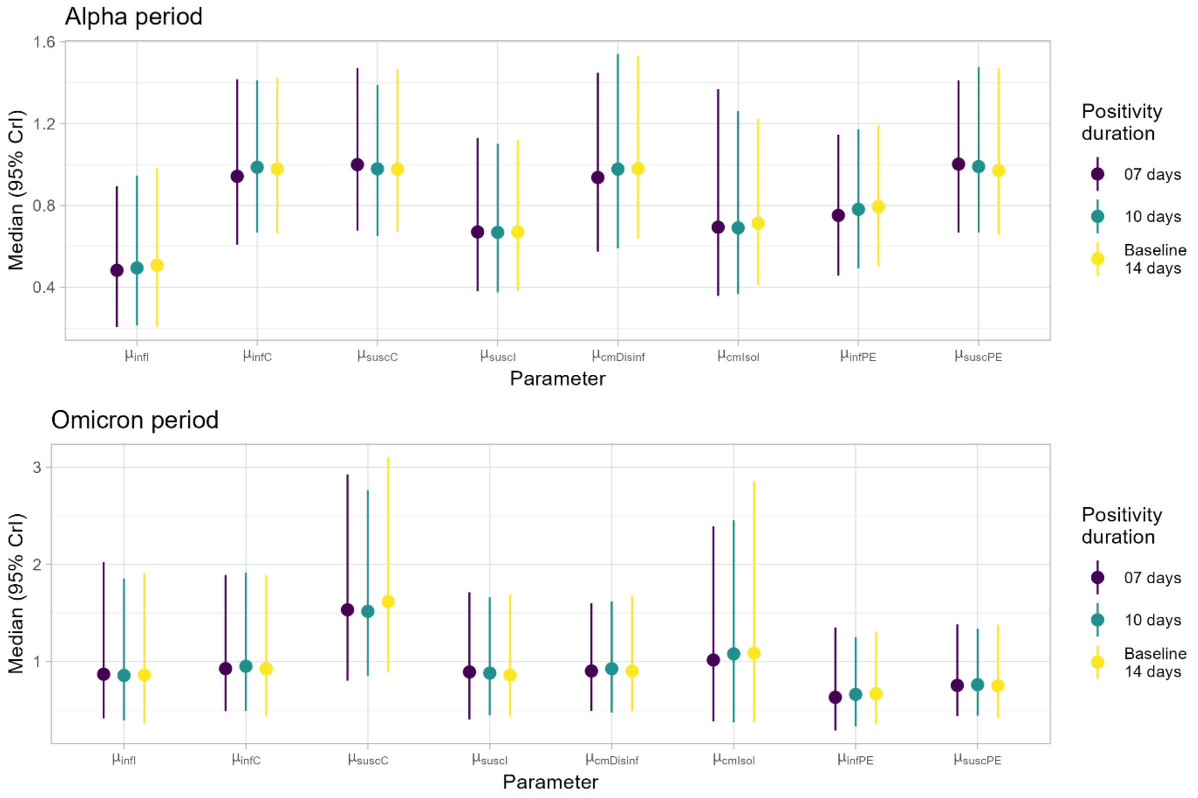

***Supplementary Figure 13. Results of the sensitivity analysis on test positivity duration: median estimates of infectiousness and susceptibility parameters for 7, 10 and 15 days of test positivity.***

#### C.8. Statistical framework

##### C.8.a. Convergence assessment

Markov chain Monte Carlo with Metropolis-Hastings algorithm was used for parameters estimation. A normal proposal was used for the parameter of dependance of transmission risk on household size $\delta$, and lognormal proposal was used for all other non-negative parameters. At each iteration, we first update parameters one by one, then perform data augmentation of symptoms onset/1$^{st}$ positive test and infection times together for each infected individual.

Chains were run for 300,000 iterations, recording one out of 500 iterations. For posterior analyses, a burn-in period of the first 30,000 iterations was discarded. Convergence was assessed by visualizing the chains (Supplementary Figure 14, Supplementary Figure 15) and comparing prior and posterior distributions (Supplementary Figure 16, Supplementary Figure 17). Convergence stability was assessed by running 5 independent chains and ensuring that results of all chains were consistent (Supplementary Figure 18, Supplementary Figure 19).

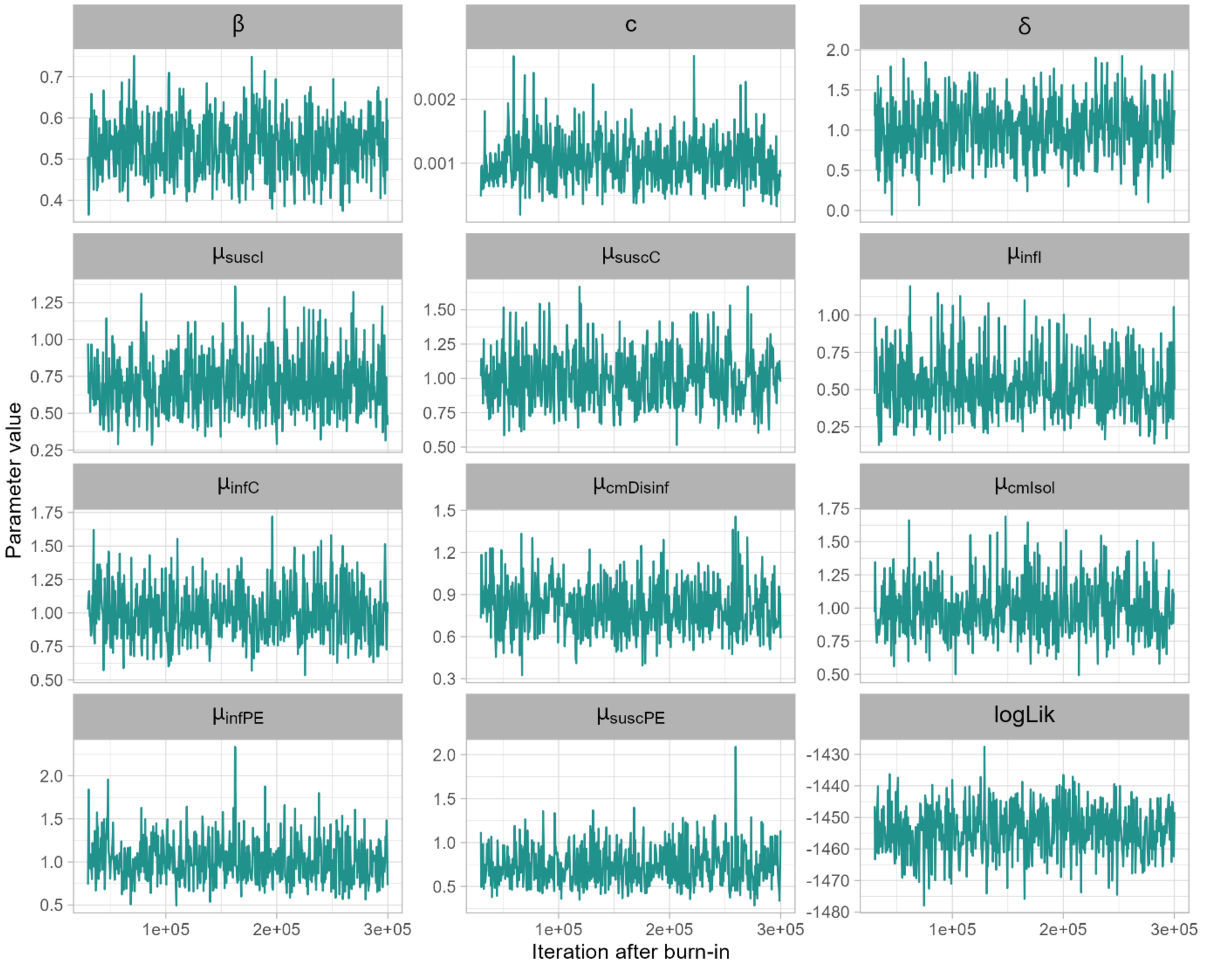

***Supplementary Figure 14. Convergence assessment (chain mixing, Alpha period). Markov chain mixing for each of the parameters estimated from the PedCovid dataset (Alpha period) using the corrected framework F1. Chains were run for 300,000 iterations, a burn-in period of 30.000 iterations was discarded.*** *$\boldsymbol{\beta}$: baseline transmission intensity, $\boldsymbol{c}$: risk of acquisition from community (/day), $\boldsymbol{\delta}$: dependence of transmission rate on household size, $\boldsymbol{\mu_{susc}}$: relative susceptibility across age (I: infant, C: child), $\boldsymbol{\mu_{inf}}$: relative infectiousness across age (I: infant, C: child), $\boldsymbol{\mu_{cm}}$: relative susceptibility across control measures (Disinf: disinfection of surfaces, Isol: isolation), $\boldsymbol{\mu_{infPE}}$ and $\boldsymbol{\mu_{suscPE}}$: relative infectiousness and susceptibility according to past exposure, $\boldsymbol{logLik}$: value of the total log-likelihood.*

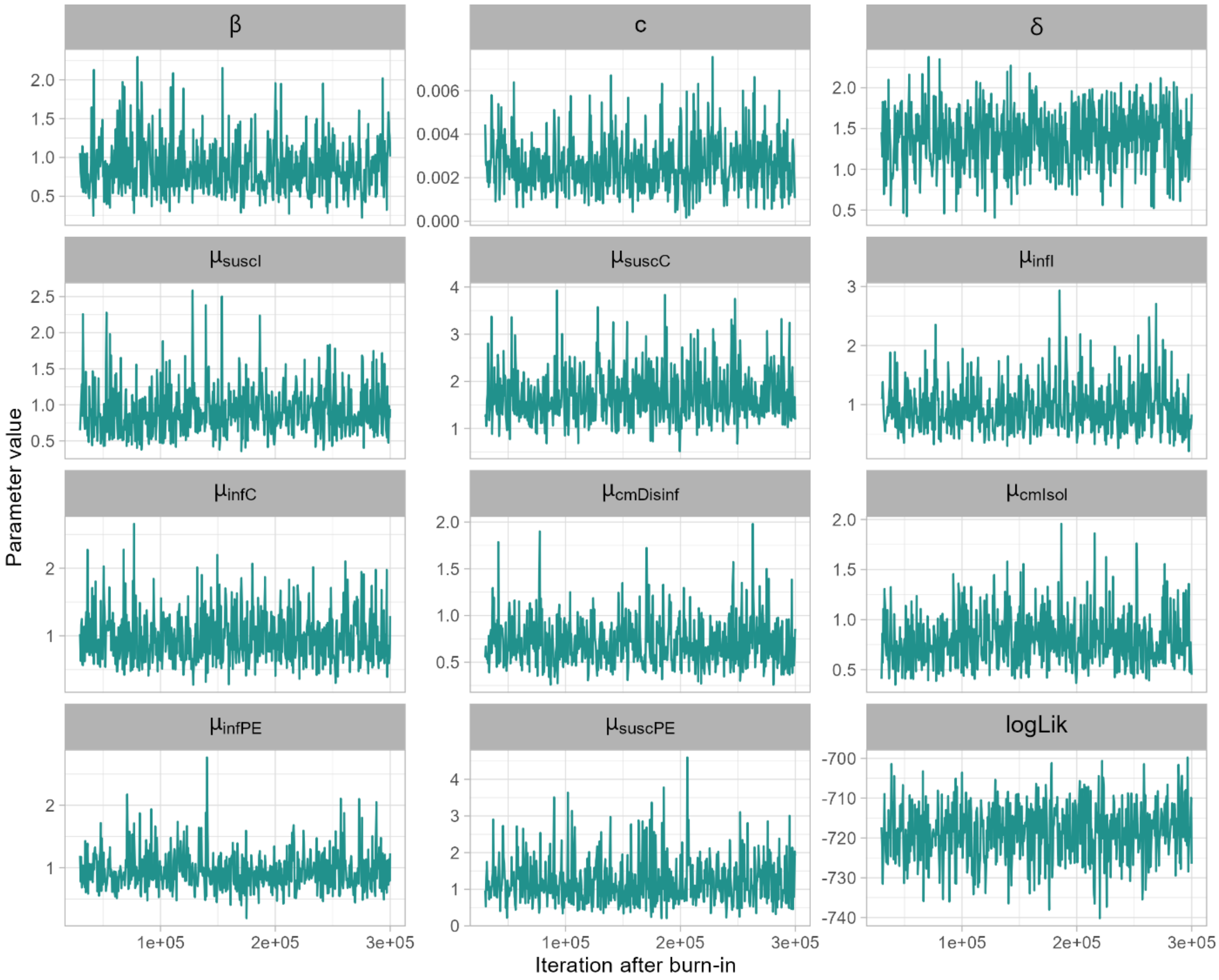

***Supplementary Figure 15. Convergence assessment (chain mixing, Omicron period). Markov chain mixing for each of the parameters estimated from the PedCovid dataset (Omicron period) using the corrected framework F1. Chains were run for 300,000 iterations, a burn-in period of 30.000 iterations was discarded.*** *$\boldsymbol{\beta}$: baseline transmission intensity, $\boldsymbol{c}$: risk of acquisition from community (/day), $\boldsymbol{\delta}$: dependence of transmission rate on household size, $\boldsymbol{\mu_{susc}}$: relative susceptibility across age (I: infant, C: child), $\boldsymbol{\mu_{inf}}$: relative infectiousness across age (I: infant, C: child), $\boldsymbol{\mu_{cm}}$: relative susceptibility across control measures (Disinf: disinfection of surfaces, Isol: isolation), $\boldsymbol{\mu_{infPE}}$ and $\boldsymbol{\mu_{suscPE}}$: relative infectiousness and susceptibility according to past exposure, $\boldsymbol{logLik}$: value of the total log-likelihood.*

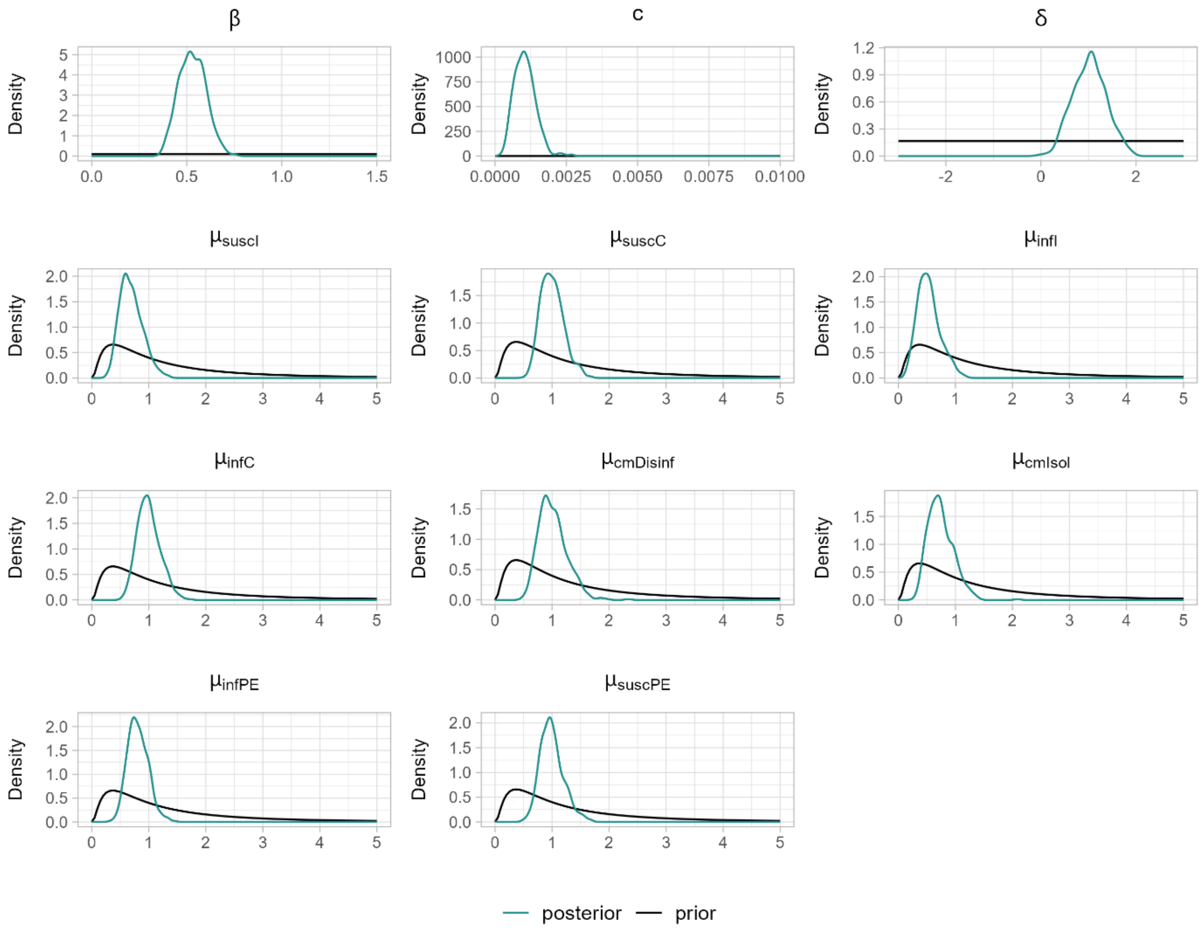

***Supplementary Figure 16. Convergence assessment (prior-posterior comparison, Alpha period). Prior and posterior distributions for all parameters after 300.000 iterations and a discarded burn-in period of 30.000 iterations (Alpha period).*** ***$\beta$***: *baseline transmission intensity,* ***$c$***: *risk of acquisition from community (/day),* ***$\delta$***: *dependence of transmission rate on household size,* ***$\mu_{susc}$***: *relative susceptibility across age (I: infant, C: child),* ***$\mu_{inf}$***: *relative infectiousness across age (I: infant, C: child),* ***$\mu_{cm}$***: *relative susceptibility across control measures (Disinf: disinfection of surfaces, Isol: isolation),* ***$\mu_{infPE}$*** *and* ***$\mu_{suscPE}$***: *relative infectiousness and susceptibility according to past exposure.*

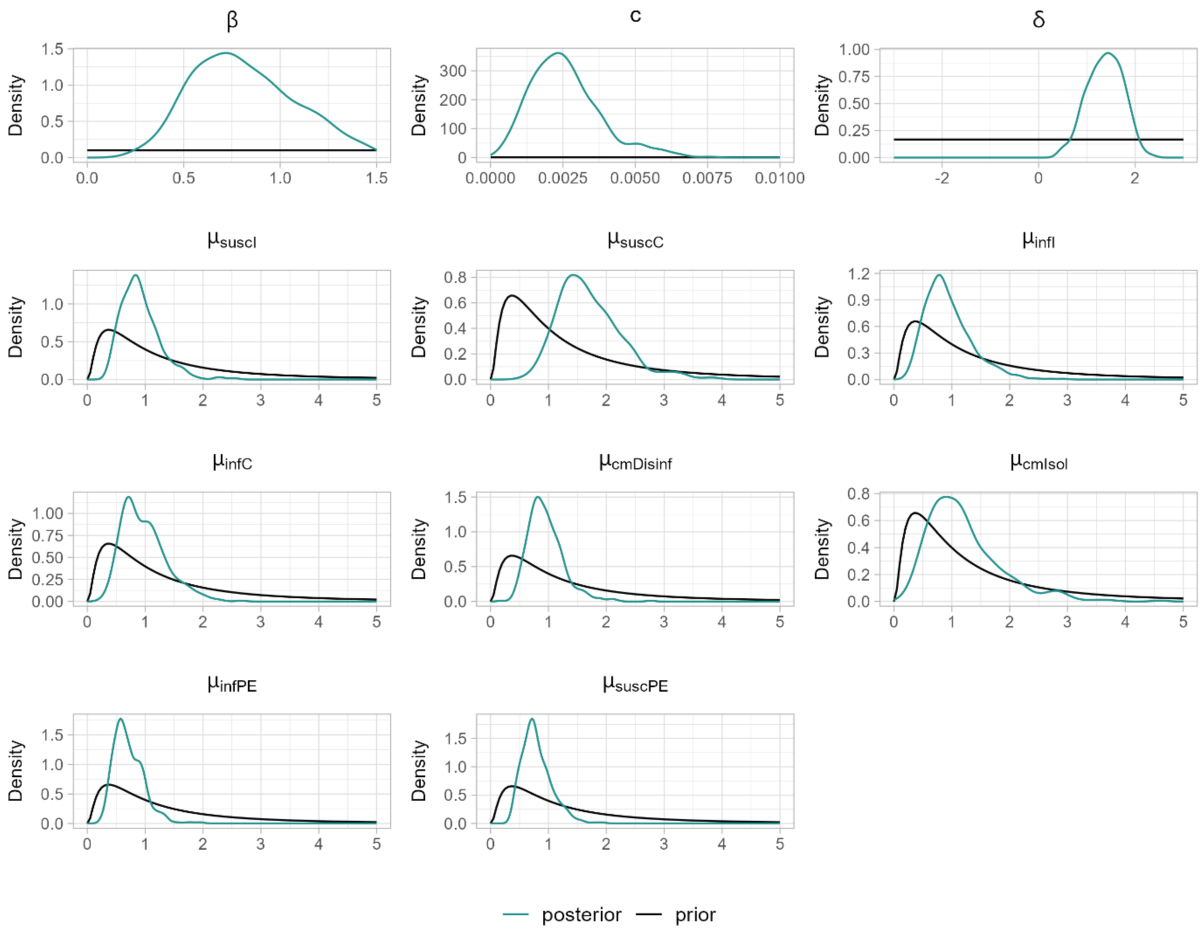

***Supplementary Figure 17. Convergence assessment (prior-posterior comparison, Omicron period). Prior and posterior distributions for all parameters after 300.000 iterations and a discarded burn-in period of 30.000 iterations (Omicron period).*** *$\boldsymbol{\beta}$: baseline transmission intensity, $\boldsymbol{c}$: risk of acquisition from community (/day), $\boldsymbol{\delta}$: dependence of transmission rate on household size, $\boldsymbol{\mu_{susc}}$: relative susceptibility across age (I: infant, C: child), $\boldsymbol{\mu_{inf}}$: relative infectiousness across age (I: infant, C: child), $\boldsymbol{\mu_{cm}}$: relative susceptibility across control measures (Disinf: disinfection of surfaces, Isol: isolation), $\boldsymbol{\mu_{infPE}}$ and $\boldsymbol{\mu_{suscPE}}$: relative infectiousness and susceptibility according to past exposure.*

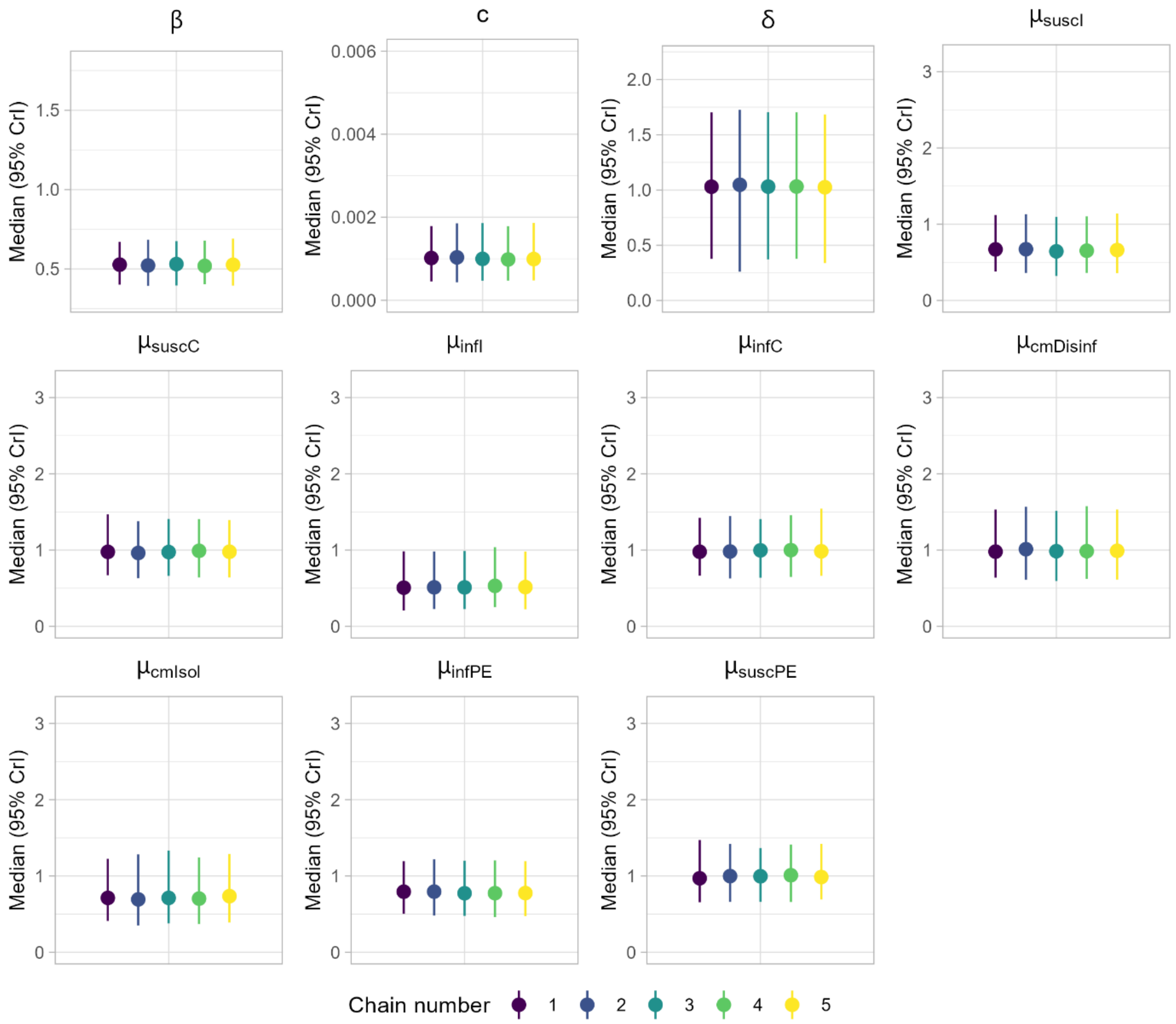

***Supplementary Figure 18. Stability assessment (Alpha period). Posterior distribution (median and 95% credible interval) for 5 independent Markov chains for each of the parameters estimated from the PedCovid dataset (Alpha period) using the corrected framework F1.*** *$\boldsymbol{\beta}$: baseline transmission intensity, **c**: risk of acquisition from community (/day), $\boldsymbol{\delta}$: dependence of transmission rate on household size, $\boldsymbol{\mu_{susc}}$: relative susceptibility across age (I: infant, C: child), $\boldsymbol{\mu_{inf}}$: relative infectiousness across age groups (I: infant, C: child), $\boldsymbol{\mu_{cm}}$: relative susceptibility across control measures (Disinf: disinfection of surfaces, Isol: isolation), $\boldsymbol{\mu_{infPE}}$ and $\boldsymbol{\mu_{suscPE}}$: relative infectiousness and susceptibility according to past exposures.*

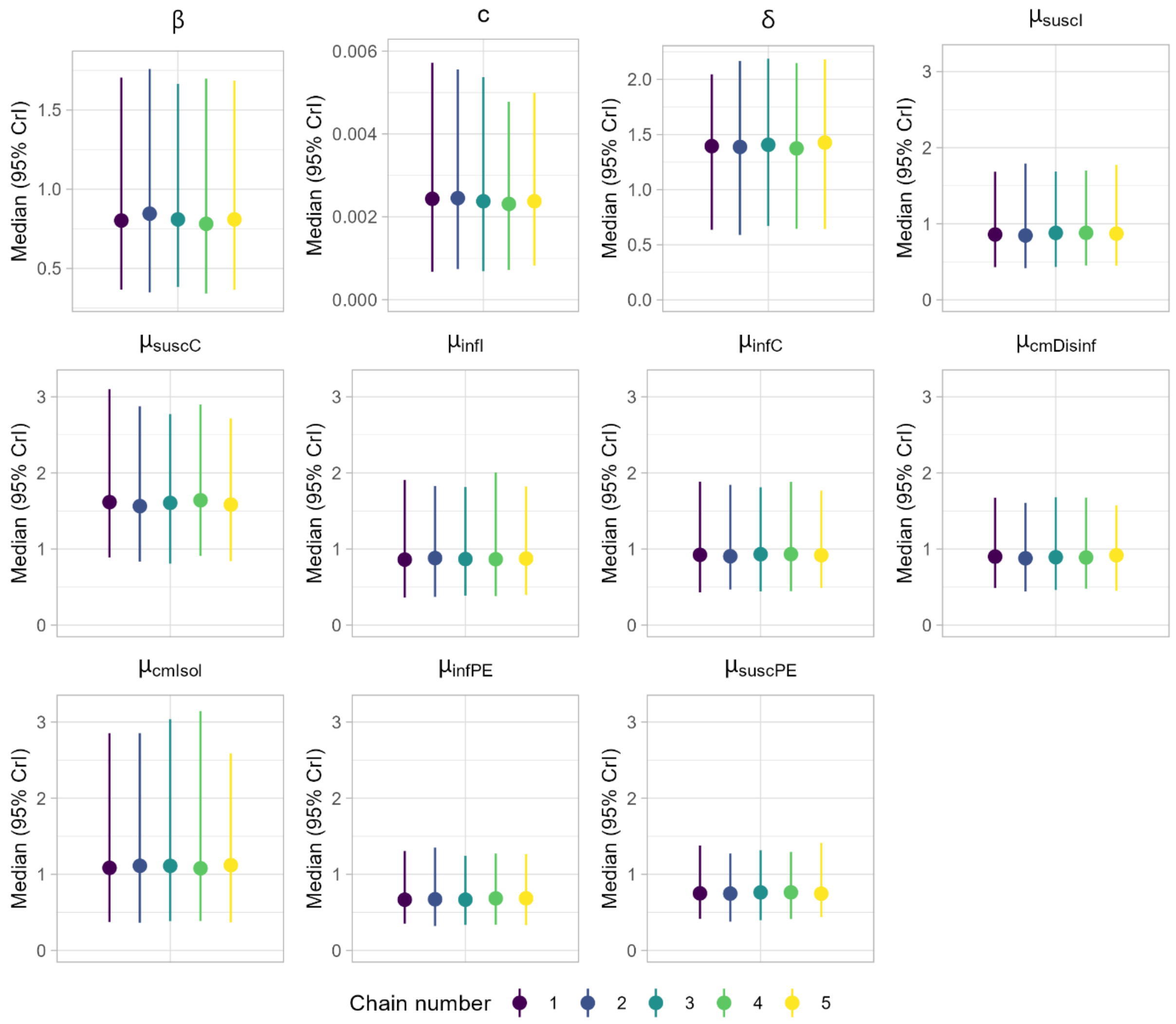

***Supplementary Figure 19. Stability assessment (Omicron period). Posterior distribution (median and 95% credible interval) for 5 independent Markov chains for each of the parameters estimated from the PedCovid dataset (Omicron period) using the corrected framework F1.*** *$\boldsymbol{\beta}$: baseline transmission intensity, $\boldsymbol{c}$: risk of acquisition from community (/day), $\boldsymbol{\delta}$: dependence of transmission rate on household size, $\boldsymbol{\mu_{susc}}$: relative susceptibility across age (I: infant, C: child), $\boldsymbol{\mu_{inf}}$: relative infectiousness across age groups (I: infant, C: child), $\boldsymbol{\mu_{cm}}$: relative susceptibility across control measures (Disinf: disinfection of surfaces, Isol: isolation), $\boldsymbol{\mu_{infPE}}$ and $\boldsymbol{\mu_{suscPE}}$: relative infectiousness and susceptibility according to past exposures.*

##### C.8.b. Sensitivity analysis on prior distributions

Prior distributions used in the MCMC are given in table Supplementary Table 2. For our main analysis, we used logNormal prior distributions for all infectiousness and susceptibility parameters. In a sensitivity analysis, we checked the impact of this choice on the estimations. We explored logNormal distributions with varied standard deviation for all infectiousness and susceptibility parameters (Supplementary Table 8). Results are given in Supplementary Figure 20. Results are not sensitive to the choice of prior distribution.

| | Distribution | Mean | SD |
|---|---|---|---|
| **Baseline** | logNormal(0,1) | 1.6 | 2.2 |
| **Narrower** | logNormal(0,0.7) | 1.4 | 1.4 |
| **Wider** | logNormal(0,2) | 2.7 | 6.9 |

***Supplementary Table 8. Distributions of the prior distributions of all infectiousness and susceptibility parameters for the sensitivity analysis.***

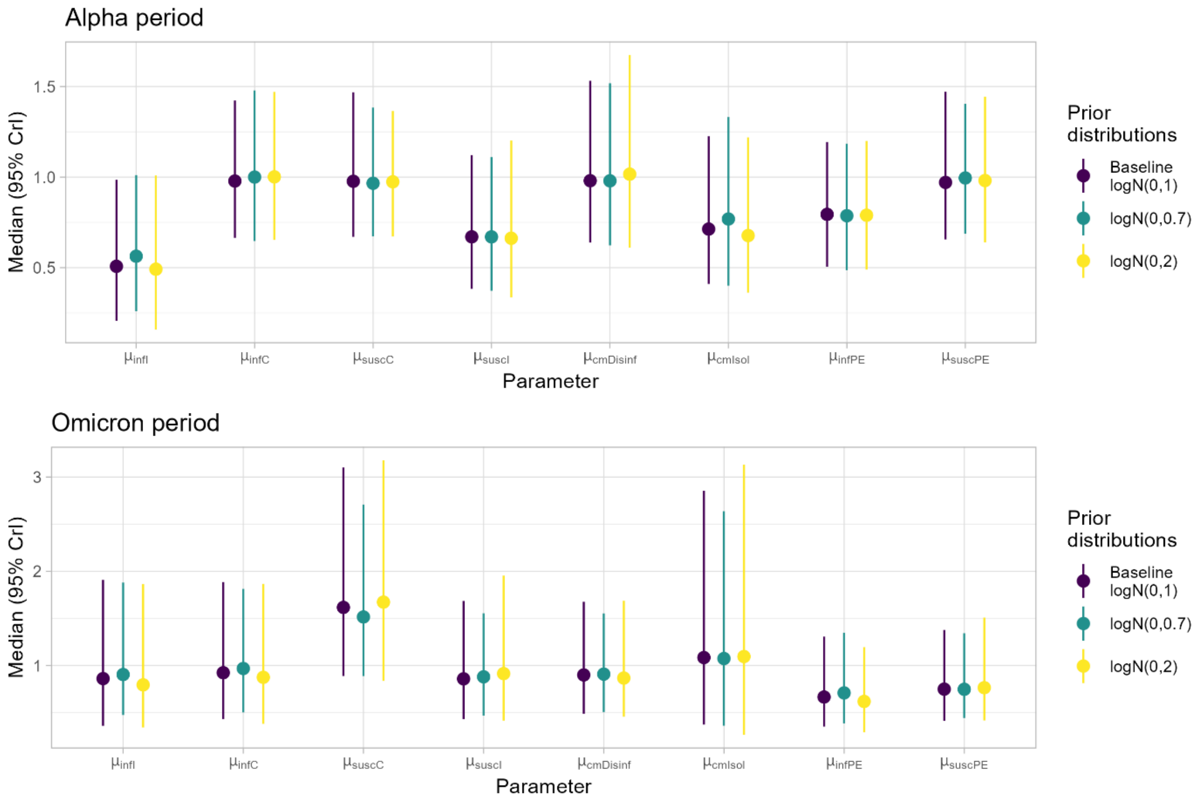

***Supplementary Figure 20. Results of the sensitivity analysis on prior distributions. Median estimates of infectiousness and susceptibility parameters for 3 logNormal distributions of varied standard deviation.*** *$\boldsymbol{\beta}$: baseline transmission intensity, $\boldsymbol{c}$: risk of acquisition from community (/day), $\boldsymbol{\delta}$: dependence of transmission rate on household size, $\boldsymbol{\mu_{susc}}$: relative susceptibility across age (I: infant, C: child), $\boldsymbol{\mu_{inf}}$: relative infectiousness across age (I: infant, C: child), $\boldsymbol{\mu_{cm}}$: relative susceptibility across control measures (Disinf: disinfection of surfaces, Isol: isolation), $\boldsymbol{\mu_{infPE}}$ and $\boldsymbol{\mu_{suscPE}}$: relative infectiousness and susceptibility according to past exposure.*

#### C.9. Reconstruction of transmission trees

For each household of the PedCovid study, we used the posterior samples of model parameters and augmented data obtained with the corrected framework (F1) to compute the probability of each transmission pair at each MCMC iteration. For each infected member, we identified the most probable infector at that iteration. The household transmission tree was then reconstructed by assigning to each infected individual the infector that was most frequently identified as the most probable across all MCMC iterations (Supplementary Figure 21).

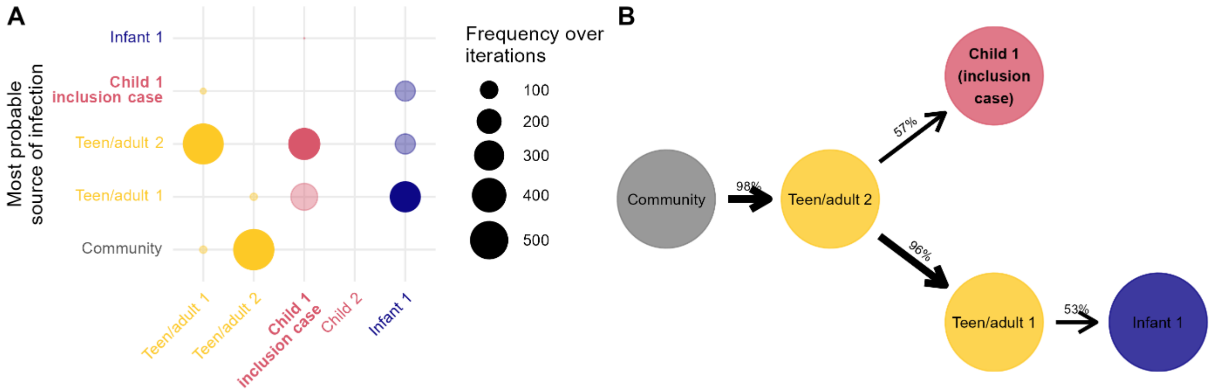

***Supplementary Figure 21. Transmissions reconstructed from the posterior distributions. A – Example of reconstruction of the most probable source of infection for each individual of household 141 over all iterations. Solid points show the most frequent transmission pair over iterations. B – Example of reconstruction of the transmission tree of household 141 using the most frequent most probable infector for each member.***
